# COVID-19 illness severity and 2-year prevalence of physical symptoms: an observational study in Iceland, Sweden, Norway and Denmark

**DOI:** 10.1101/2023.04.18.23288720

**Authors:** Qing Shen, Emily E. Joyce, Omid V. Ebrahimi, Maria Didriksen, Anikó Lovik, Karen Sól Sævarsdóttir, Ingibjörg Magnúsdóttir, Dorte Helenius Mikkelsen, Anna Bára Unnarsdóttir, Arna Hauksdóttir, Asle Hoffart, Anna K. Kähler, Edda Björk Thórdardóttir, Elías Eythórsson, Emma M. Frans, Gunnar Tómasson, Helga Ask, Hrönn Hardardóttir, Jóhanna Jakobsdóttir, Kelli Lehto, Li Lu, Ole A. Andreassen, Patrick F. Sullivan, Runólfur Pálsson, Christian Erikstrup, Sisse Rye Ostrowski, Thomas Werge, Thor Aspelund, Ole B. V. Pedersen, Sverre Urnes Johnson, Fang Fang, Unnur Anna Valdimarsdóttir

**Affiliations:** Clinical Research Center for Mental Disorders, Shanghai Pudong New Area Mental Health Center, Tongji University School of Medicine, Shanghai, China; Institute for Advanced Study, Tongji University, Shanghai, China; Centre of Public Health Sciences, Faculty of Medicine, School of Health Sciences, University of Iceland, Reykjavik, Iceland; Unit of Integrative Epidemiology, Institute of Environmental Medicine, Karolinska Institutet, Stockholm, Sweden; Department of Psychology, University of Oslo, Oslo, Norway; Modum Bad Psychiatric Hospital and Research Center, Vikersund, Norway; Department of Psychiatry, University of Oxford, Oxford, United Kingdom; Oxford Precision Psychiatry Lab, NIHR Oxford Health Biomedical Research Centre, Oxford, United Kingdom; Department of Clinical Immunology, Copenhagen University Hospital, Rigshospitalet, Copenhagen, Denmark; Methodology and Statistics Unit, Institute of Psychology, Leiden University, Leiden, The Netherlands; Institute of Biological Psychiatry, Mental Health Services, Copenhagen University Hospital, Copenhagen, Denmark; The Lundbeck Foundation Initiative for Integrative Psychiatric Research, Copenhagen and Aarhus, Denmark; Department of Medical Epidemiology and Biostatistics, Karolinska Institutet, Stockholm, Sweden; Mental Health Services, Landspitali, The National University Hospital of Iceland, Reykjavik, Iceland; Internal Medicine and Emergency Services, Landspitali, The National University Hospital of Iceland, Reykjavik, Iceland; Department of Mental Disorders, Norwegian Institute of Public Health, Oslo, Norway; Estonian Genome Centre, Institute of Genomics, University of Tartu, Estonia; NORMENT Centre, Institute of Clinical Medicine, University of Oslo, Oslo, Norway; NORMENT Centre, Division of Mental Health and Addiction, Oslo University Hospital, Oslo, Norway; Departments of Genetics and Psychiatry, University of North Carolina, Chapel Hill, North Carolina, USA; Faculty of Medicine, School of Health Sciences, University of Iceland, Reykjavik, Iceland; Department of Clinical Medicine, Faculty of Health and Medical Sciences, University of Copenhagen, Copenhagen, Denmark; Lundbeck Foundation Center for GeoGenetics, GLOBE Institute, University of Copenhagen, Copenhagen, Denmark; The Icelandic Heart Association, Kopavogur, Iceland; Department of Clinical Immunology, Zealand University Hospital, Denmark; Department of Epidemiology, Harvard T.H. Chan School of Public Health, Boston, Massachusetts, USA

**Keywords:** physical symptom, long covid, cohort, COVID-19

## Abstract

**Background:** Persistence of physical symptoms after SARS-CoV-2 infection is a major public health concern, although evidence from large observational studies remain scarce. We aimed to assess the prevalence of physical symptoms in relation to acute illness severity up to more than 2-years after diagnosis of COVID-19.

**Methods:** This multinational study included 64 880 adult participants from Iceland, Sweden, Denmark, and Norway with self-reported data on COVID-19 and physical symptoms from April 2020 to August 2022. We compared the prevalence of 15 physical symptoms, measured by the Patient Health Questionnaire (PHQ-15), among individuals with or without a confirmed COVID-19 diagnosis, by acute illness severity, and by time since diagnosis. We additionally assessed the change in symptoms in a subset of Swedish adults with repeated measures, before and after COVID-19 diagnosis.

**Findings:** During up to 27 months of follow-up, 22 382 participants (34.5%) were diagnosed with COVID-19. Individuals who were diagnosed with COVID-19, compared to those not diagnosed, had an overall 37% higher prevalence of severe physical symptom burden (PHQ-15 score ≥ 15, adjusted prevalence ratio [PR] 1.37 [95% confidence interval [CI] 1.23-1.52]). The prevalence was associated with acute COVID-19 severity: individuals bedridden for seven days or longer presented with the highest prevalence (PR 2.25[1.85-2.74]), while individuals never bedridden presented with similar prevalence as individuals not diagnosed with COVID-19 (PR 0.92 [0.68-1.24]). The prevalence was statistically significantly elevated among individuals diagnosed with COVID-19 for eight of the fifteen measured symptoms: shortness of breath, chest pain, dizziness, heart racing, headaches, low energy/fatigue, trouble sleeping, and back pain. The analysis of repeated measurements rendered similar results as the main analysis.

**Interpretation:** These data suggest an elevated prevalence of some, but not all, physical symptoms during up to more than 2 years after diagnosis of COVID-19, particularly among individuals suffering a severe acute illness.

**Funding:** This work was mainly supported by grants from NordForsk (COVIDMENT, grant number 105668 and 138929) and Horizon2020 (CoMorMent, 847776). See Acknowledgements for further details on funding.

**Research in context:** *Evidence before the study:* As the majority of the global population has contracted COVID-19, persistence of physical symptoms after SARS-CoV-2 infection (*Long* COVID or post COVID-19 condition) has become a major public health concern. We searched PubMed for studies assessing physical symptoms after COVID-19, published by March 22, 2023. The search term was (physical symptoms after covid) AND LitCLONGCOVID [Pubmed filter]. We reviewed 82 studies, after excluding those not on humans or not published in English. High prevalence of multiple physical symptoms, mainly fatigue, shortness of breath, headache, muscle and chest pain, has been reported, mostly based on small samples of hospitalized patients confined to three to six months after diagnosis. A comprehensive assessment of long-term prevalence of physical symptoms beyond six months after diagnosis and among non-hospitalized patients is lacking.

*Added value of this study:* We included 64 880 participants from the general population of four Nordic countries, of whom 22 382 had been diagnosed with COVID-19 up to 2 years earlier (<1% hospitalized due to COVID-19). Individuals diagnosed with COVID-19 reported a 37% higher prevalence of overall severe physical symptom burden compared to individuals not diagnosed with COVID-19. We found that shortness of breath, chest pain, dizziness, headaches, and low energy/fatigue were particularly increased among individuals with COVID-19 diagnosis. Individuals bedridden for seven days or more during the acute illness phase (9.6% of the patients) showed the greatest and most persistent elevation in prevalence of severe physical symptoms while individuals not bedridden during the acute COVID-19 illness showed no increase in prevalence of physical symptoms compared to those not diagnosed.

*Implications of the available evidence:* Our findings provide timely and valuable evidence to demonstrate the constitution of Long COVID and the long-term health consequences after recovery from COVID-19 in the general population. The long-term risk of severe physical symptom burden is distinctly associated with acute illness severity, highlighting the importance of sustained monitoring of physical symptoms among the group of patients who suffered severe acute illness course.

## Introduction

The COVID-19 pandemic continues to have substantial public health consequences three years after its outbreak in 2020. As of March 2023, more than 683 million individuals worldwide have been infected by the SARS-CoV-2 virus^1^ and an estimated 10-20% of these individuals will continue to experience a variety of symptoms after recovery.^2^ The presence of long-term symptoms persisting beyond two months after infection has been recognized as *Long* COVID or post-COVID-19 condition.^3,4^ In addition to flu-like symptoms, studies suggest that other physical symptoms can extend beyond the disease course,^4,5^ including fatigue,^3–6^ shortness of breath,^4^ headache,^5,6^ and myalgias and chest pain.^3,4,6^ Most of these studies are based on small patient samples,^7,8^ with three to six month follow-up post-diagnosis and, notably, focused on hospitalized patients.^5^ General population data on the prevalence of physical symptoms after COVID-19 diagnosis over a longer period of time is lacking, especially by measures of acute illness severity.^5,6^ Importantly, the absence of comparison to populations without a confirmed COVID-19 diagnosis has largely limited the interpretation of these findings.^4,6,8^ Also, the use of validated instruments to quantify physical symptom severity is needed because assessment of individual physical symptoms has varied greatly in the literature,^6^ making it difficult for direct comparisons.

With this background, we leveraged data from four *Nordic* cohorts of the COVIDMENT Consortium^9^ to investigate the prevalence of physical symptom burden up to 27 months following a COVID-19 diagnosis with a focus on analysing the severity of acute COVID-19 illness and time since diagnosis. We further utilized repeated measures of the physical symptoms to assess the change in the prevalence of physical symptoms before and after COVID-19 diagnosis in a subset of adult study participants in Sweden.

## Methods

### Study population and design

We included four cohorts from the COVIDMENT Consortium with data collection on physical symptoms: The Icelandic COVID-19 National Resilience Cohort (C-19 Resilience, N=14 358, from April 2020 to August 2021), The Swedish Omtanke2020 Study (N=18 190, from July 2021 to February 2022), The Norwegian COVID-19, Mental Health and Adherence Project (MAP-19, N=3 310, March 2022), and The Danish Blood Donor Study (DBDS, N=29 958, from June to August 2022).^9^ All study participants provided electronic informed consent. Ethical approvals were obtained for each participating cohort from national or regional bioethics committees (Supplementary Table S1). After exclusion of study participants with incomplete information on COVID-19 diagnosis and physical symptoms, 64 880 individuals were included for further analysis (Supplementary Figure S1 flowchart).

### Data collection and assessment

COVID-19 diagnosis was assessed by self-reported SARS-CoV-2 infection from a positive RT-PCR test. Participants who reported a confirmed diagnosis of COVID-19 during the study period were referred to as the COVID group whereas the remaining participants were referred to as the non-COVID group. Time since diagnosis was defined as the time interval between the reported date of diagnosis and physical symptom data collection, and was coded as 0-2 months, 3-5 months, 6-9 months, and 10-27 months (up to 16 months in C-19 Resilience, up to 22 months in Omtanke2020, up to 24 months in MAP-19, and up to 27 months in DBDS). The illness severity during the acute phase of COVID-19 was determined by self-reported number of days confined to bed (not bedridden, bedridden 1-6 days, or bedridden 7 days or longer), and hospitalization due to COVID-19 infection (yes or no).^10^

We used the 15-item Patient Health Questionnaire (PHQ-15) to measure the severity of physical symptoms most commonly recognized in outpatient settings. The PHQ-15 is a widely used instrument and has been validated in different populations.^11^ Each symptom is scored as 0 (“not bothered at all”), 1 (“bothered a little”), or 2 (“bothered a lot”).^12^ Consistent with previous studies, a cut-off of PHQ-15 score≥15 was defined as experiencing *severe physical symptom burden* (termed as severe symptoms) in our study, as this score indicates severe symptomology.^12,13^ We used multiple imputation to estimate the individual physical symptom responses for participants with less than 25% missingness in the PHQ-15.^13,14^ Because data on physical symptoms was collected prospectively, we used assessments since enrollment for the non-COVID group, and since diagnosis for the COVID group.

Several covariates were included in the analysis, when available (Supplementary Table S1). The included covariates were age (discrete numeric in years), gender (male, female, or other), average monthly income (low, low-medium, medium, medium-high, or high income; not available in Omtanke2020), residency (capital or elsewhere), relationship status (single or in a relationship; not available in DBDS), body mass index (BMI, categorized as <25 kg/m^2^ [normal or underweight], 25-30 kg/m^2^ [overweight], or >30 kg/m^2^ [obese]), current smoking (no or yes), habitual drinking (no or yes), history of psychiatric disorder (no or yes), pre-existing somatic comorbidity (no, one or more comorbidities), and response period (June 2020 or earlier, July-September 2020, October-December 2020, January-March 2021, April-June 2021, July-September 2021, October-December 2021, or January 2022 or after). We also included variables of current mental health status measured at the same survey (no or yes) for potential depression (Patient Health Questionnaire (PHQ-9)), anxiety (General Anxiety Disorder (GAD-7)) and COVID-19-related distress symptoms (Primary Care PTSD Screen for DSM-5 (PC-PTSD-5) and the PTSD checklist for DSM-5; not available in DBDS), as described in our previous study.^10^

### Statistical analysis

We first described the distribution of the above covariates between the COVID group and non-COVID group, as well as by acute illness severity and time since diagnosis for the COVID-19 group. We conducted a cross-sectional analysis comparing the prevalence of severe symptoms (PHQ-15 ≥ 15) in the COVID group, overall and by illness severity (bedridden and hospitalization) and time since diagnosis, to that of the non-COVID group. We applied a robust (modified) Poisson regression model to estimate prevalence ratios (PRs) with 95% confidence intervals (CIs), with a quasi-likelihood model used to fit a binary outcome.^15^ A sandwich estimator with exchangeable working correlation structure was applied in the model to control for intra-individual correlation when repeated measures were available.^16^ The PRs were calculated in crude models and then in adjusted models controlling for age, gender, average monthly income, residency, relationship status, BMI, current smoking, habitual drinking, previous diagnosis of psychiatric disorder, pre-existing somatic comorbidity, and response period. Covariates not measured in an individual cohort were not adjusted for in the analysis of that cohort. All analyses were performed among individuals with complete information on variables. We also stratified by gender, age, mental health indicators, and pre-existing somatic comorbidity to investigate if these factors would modify the association between COVID-19 and severe symptoms. To investigate of the associations for individual symptoms, we estimated PRs for the 15 measured symptoms individually, analysing separately reporting being “bothered a little” or being “bothered a lot”. The analysis of PRs for reporting “bothered a lot” was further performed by illness severity and by time since diagnosis.

In the Swedish Omtanke2020 Study, a subset of individuals with a COVID-19 diagnosis reported repeated measures of physical symptoms: one *prior to* and one following their COVID-19 diagnosis. We conducted a pairwise analysis assessing prevalence of individual symptoms comparing post-COVID (first response after diagnosis) to pre-COVID (last response before diagnosis) measures. The extension of the modified Poisson regression models mentioned above was performed to assess the difference in prevalence of individual symptoms between the two time-points.

We performed the above analyses using a standardized analysis protocol in all four cohorts, and finally, to combine aggregated data from the cohorts, we meta-analysed the output from each cohort with a random-effects model using the metafor package in R to estimate an overall result for all analyses.^17^ Heterogeneity of the findings was examined using the *I*^2^ statistic.^18^ Statistical analyses were conducted in R (version 4.0.5). This study is reported according to the Strengthening the Reporting of Observational studies in Epidemiology (STROBE) checklist.

## Results

### Baseline characteristics

Of the 64 880 participants, 22 382 (34.5%) reported having been diagnosed with COVID-19. Persons with COVID-19 were younger, had a lower BMI, and had a lower proportion of pre-existing psychiatric disorder and somatic comorbidity, compared with the non-COVID group (Table 1). The demographics varied across the cohorts, with participants of MAP-19 being younger in age and more likely to be single than participants of other cohorts (Supplementary Table S2). Among persons with COVID-19, 28.0% were bedridden during acute infection (18.4% for 1-6 days; and 9.6% for 7 days or more) and 1.0% were hospitalized (Table 1).

**Table 1.** Background characteristics of 64 880 study participants *NOT* diagnosed or diagnosed with COVID-19.

|  | Individuals *NOT* diagnosed<br>with COVID-19<br>N (%) | Individuals diagnosed<br>with COVID-19<br>N (%) |
| --- | --- | --- |
| **Total** | 42 498 | 22 382 |
| C-19 Resilience (IS) | 13 284 (31.0%) | 1 074 (4.7%) |
| Omtanke2020 (SE) | 14 841 (34.7%) | 3 349 (14.5%) |
| MAP-19 (NO) | 1 701 (4.0%) | 1 609 (7.0%) |
| DBDS (DK) | 12 672 (29.6%) | 16 350 (71.0%) |
| **Gender** |  |  |
| Male | 13 290 (31.3%) | 8 507 (38.0%) |
| Female | 29 114 (68.5%) | 13 856 (61.9%) |
| Other | 80 (0.2%) | 13 (0.1%) |
| Missing | 14 (<0.1%) | 6 (<0.1%) |
| **Age** |  |  |
| Mean. years (SD) | 54 (14.7) | 49 (11.7) |
| 18-29 years | 3 334 (7.8%) | 2 617 (11.7%) |
| 30-39 years | 5 117 (12.0%) | 3 771 (16.8%) |
| 40-49 years | 6 776 (15.9%) | 4 696 (21.0%) |
| 50-59 years | 9 979 (23.5%) | 5 510 (24.6%) |
| 60-69 years | 10 309 (24.3%) | 4 005 (17.9%) |
| 70 years or older | 6 968 (16.4%) | 1 776 (7.9%) |
| Missing | 14 (<0.1%) | 7 (<0.1%) |
| **Average monthly income** |  |  |
| Low income | 5 499 (12.9%) | 2 786 (12.4%) |
| Low-medium income | 6 261 (14.7%) | 3 815 (17.0%) |
| Medium income | 6 362 (15.0%) | 4 463 (19.9%) |
| Medium-high income | 4 731 (11.1%) | 3 605 (16.1%) |
| High income | 3 606 (8.5%) | 3 848 (17.2%) |
| Missing | 1 198 (2.8%) | 516 (2.3%) |
| Not measured<sup>a</sup> | 14 841 (34.9%) | 3 349 (15.0%) |
| **Residence<sup>b</sup>** |  |  |
| Capital area | 21 544 (50.7%) | 9 752 (43.6%) |
| Elsewhere | 20 735 (48.8%) | 12 589 (56.2%) |
| Missing or abroad | 219 (0.5%) | 41 (0.2%) |
| **Relationship status** |  |  |
| Single | 7 925 (18.6%) | 1 593 (7.1%) |
| In a relationship | 21 660 (51.0%) | 4 380 (19.6%) |
| Missing | 241 (0.6%) | 59 (0.3%) |
| Not measured<sup>a</sup> | 12 672 (29.8%) | 16 350 (73.0%) |
| **BMI (kg/m<sup>2</sup>)** |  |  |
| < 25 Normal or low weight | 17 591 (41.4%) | 10 090 (45.1%) |
| 25-30 Overweight | 14 904 (35.1%) | 8 035 (35.9%) |
| > 30 Obese | 8 708 (20.5%) | 3 858 (17.2%) |
| Missing | 1 295 (3.0%) | 399 (1.8%) |
| **Current smoking status** |  |  |
| No | 36 848 (86.7%) | 20 279 (90.6%) |
| Yes | 5 236 (12.3%) | 2 013 (9.0%) |
| Missing | 414 (1.0%) | 90 (0.4%) |
| **Habitual drinking** |  |  |
| No | 35 455 (83.4%) | 18 584 (83.0%) |
| Yes | 7 043 (16.6%) | 3 798 (17.0%) |
| **History of psychiatric disorder** |  |  |
| No | 31 186 (73.4%) | 18 472 (82.5%) |
| Yes | 10 705 (25.2%) | 3 790 (16.9%) |
| Missing | 607 (1.4%) | 120 (0.5%) |
| **Pre-existing comorbidity** |  |  |
| No | 30 027 (70.7%) | 18 299 (81.8%) |
| Yes | 12 156 (28.6%) | 3 964 (17.7%) |
| Missing | 315 (0.7%) | 119 (0.5%) |
| **Current depressive symptoms<sup>c</sup>** |  |  |
| No | 36 174 (85.1%) | 19 397 (86.7%) |
| Yes | 5 446 (12.8%) | 2 644 (11.8%) |
| Missing | 878 (2.1%) | 341 (1.5%) |
| **Current anxiety symptoms<sup>d</sup>** |  |  |
| No | 38 805 (91.3%) | 21 146 (94.5%) |
| Yes | 3 360 (7.9%) | 1 209 (5.4%) |
| Missing | 333 (0.8%) | 27 (0.1%) |
| **Current COVID-19 related distress symptoms<sup>e</sup>** |  |  |
| No | 21 531 (50.7%) | 4 840 (21.6%) |
| Yes | 7 961 (18.7%) | 1 163 (5.2%) |
| Missing | 334 (0.8%) | 29 (0.1%) |
| Not measured<sup>a</sup> | 12 672 (29.8%) | 16 350 (73.0%) |
| **Response period** |  |  |
| June 2020 or earlier | 12 744 (30.0%) | 235 (1.0%) |
| July-September 2020 | 136 (0.3%) | 6 (0.0%) |
| October-December 2020 | 363 (0.9%) | 178 (0.8%) |
| January-March 2021 | 37 (0.1%) | 618 (2.8%) |
| April-June 2021 | 4 (0.0%) | 37 (0.2%) |
| July-September 2021 | 10 072 (23.7%) | 1 434 (6.4%) |
| October-December 2021 | 1 970 (4.6%) | 423 (1.9%) |
| January 2022 or after | 17 172 (40.4%) | 19 451 (86.9%) |
| **Illness severity – Time bedridden** |  |  |
| Not bedridden | - | 15 027 (67.1%) |
| Bedridden 1-6 days | - | 4 127 (18.4%) |
| Bedridden 7 days or more | - | 2 148 (9.6%) |
| Missing | - | 1 080 (4.8%) |
| **Illness severity – Hospitalization** |  |  |
| Not hospitalized | - | 20 670 (92.4%) |
| Hospitalized | - | 230 (1.0%) |
| Missing or not measured | - | 1 482 (6.6%) |
| **Time since COVID-19 diagnosis** |  |  |
| 0-2 months | - | 3 150 (14.1%) |
| 3-5 months | - | 10 879 (48.6%) |
| 6-9 months | - | 4 822 (21.5%) |
| >10 months | - | 3 242 (14.5%) |
| Missing | - | 289 (1.3%) |

<sup>a</sup> Average monthly income was not measured in Omtanke2020 (SE). Relationship status and current COVID-19-related distress symptoms were not measured in DBDS (DK).
<sup>b</sup> *Missing or abroad* refers to either missing in four cohorts or reported as living abroad in C-19 Resilience (IS).
<sup>c</sup> Current depressive symptoms were measured using Patient Health Questionnaire (PHQ-9), with recommended cut-off of $\geq 10$ indicating *Yes* to potentially having this condition in all cohorts.
<sup>d</sup> Current anxiety symptoms were measured using General Anxiety Disorder (GAD-7) in C-19 Resilience (IS), Omtanke2020 (SE) and MAP-19 (NO), while measured by Angst-Symptom-Spørgeskemaet (ASS) in DBDS (DK), with a recommended cut-off of $\geq 10$ indicating *Yes* to having this condition.
<sup>e</sup> Current COVID-19-related distress symptoms were measured using the modified Primary Care PTSD Screen for DSM-5 (PC-PTSD-5) with a recommended cut-off of $\geq 4$ , and the PTSD checklist for DSM-5 (PCL-5), indicating *Yes* to having this condition.

### Physical symptom burden

The prevalence of severe symptoms was higher among the COVID-19 group than the non-COVID group in all cohorts (Supplementary Table S3): 16.0% vs. 9.7% in C-19 Resilience, 8.0% vs. 5.5% in Omtanke2020, 8.5% vs. 7.6% in MAP-19, and 1.6% vs. 1.2% in DBDS. Compared with the non-COVID group, individuals diagnosed with COVID-19 had an overall higher prevalence of severe symptoms during the entire study period, in each cohort as well as combined (overall adjusted PR 1.37 [95% CI 1.23-1.52], *I*^*2*^ 43.2%; p<0.001) (Figure 1). The prevalence increase was noted regardless of gender, age groups, existence of depressive, anxiety or COVID-19 related distress symptoms, or pre-existing somatic comorbidity, although higher prevalence increase was observed for individuals without depressive or anxiety symptoms (Supplementary Figure S2).

**Figure 1.**
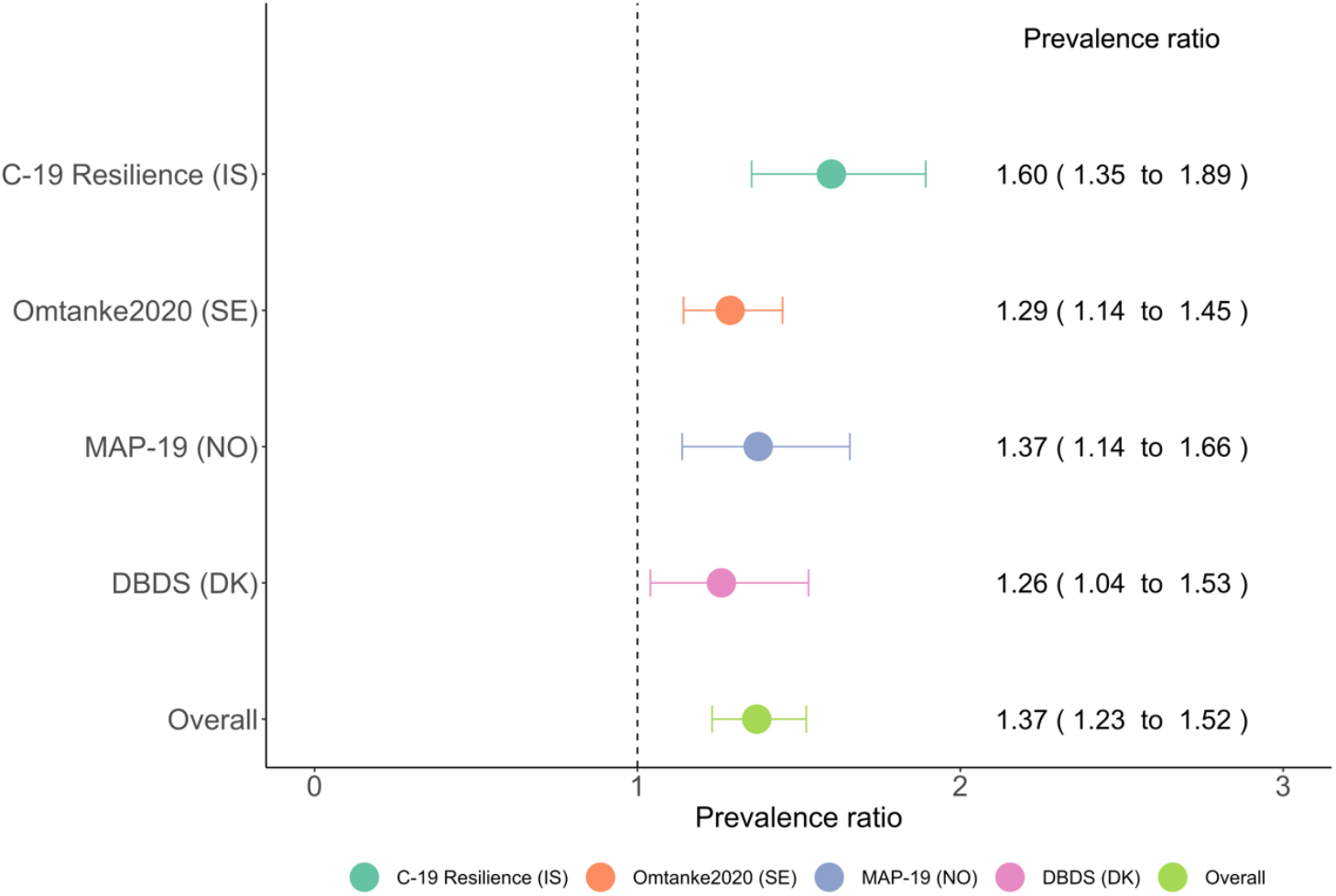
Prevalence ratio (95% confidence interval) of severe physical symptom burden (PHQ-15 ≥15) among individuals with COVID-19 compared with individuals *NOT* diagnosed with COVID-19 in the four cohorts, and a meta-analysis (*I*^*2*^ 43.2%)^a^. ^**a**^ Prevalence ratios were adjusted for age, gender, residency, average monthly income, current smoking, BMI, pre-existing comorbidity, relationship status, habitual drinking, previous diagnosis of psychiatric disorder, and response period. Income was not available in Omtanke2020 (SE), and relationship status was not available in DBDS (DK).

Longer time bedridden during acute infection was associated with a higher prevalence of severe symptoms in a dose-response manner, and the highest prevalence was consistently observed up to 27 months after diagnosis among participants bedridden for 7 days or more (combined results in Figure 2 and by cohort in Supplementary Figure S3 A). A higher prevalence was also observed among individuals hospitalized for COVID-19 from 2 months to 22 months following diagnosis (Supplementary Figure S3 B).

**Figure 2.**
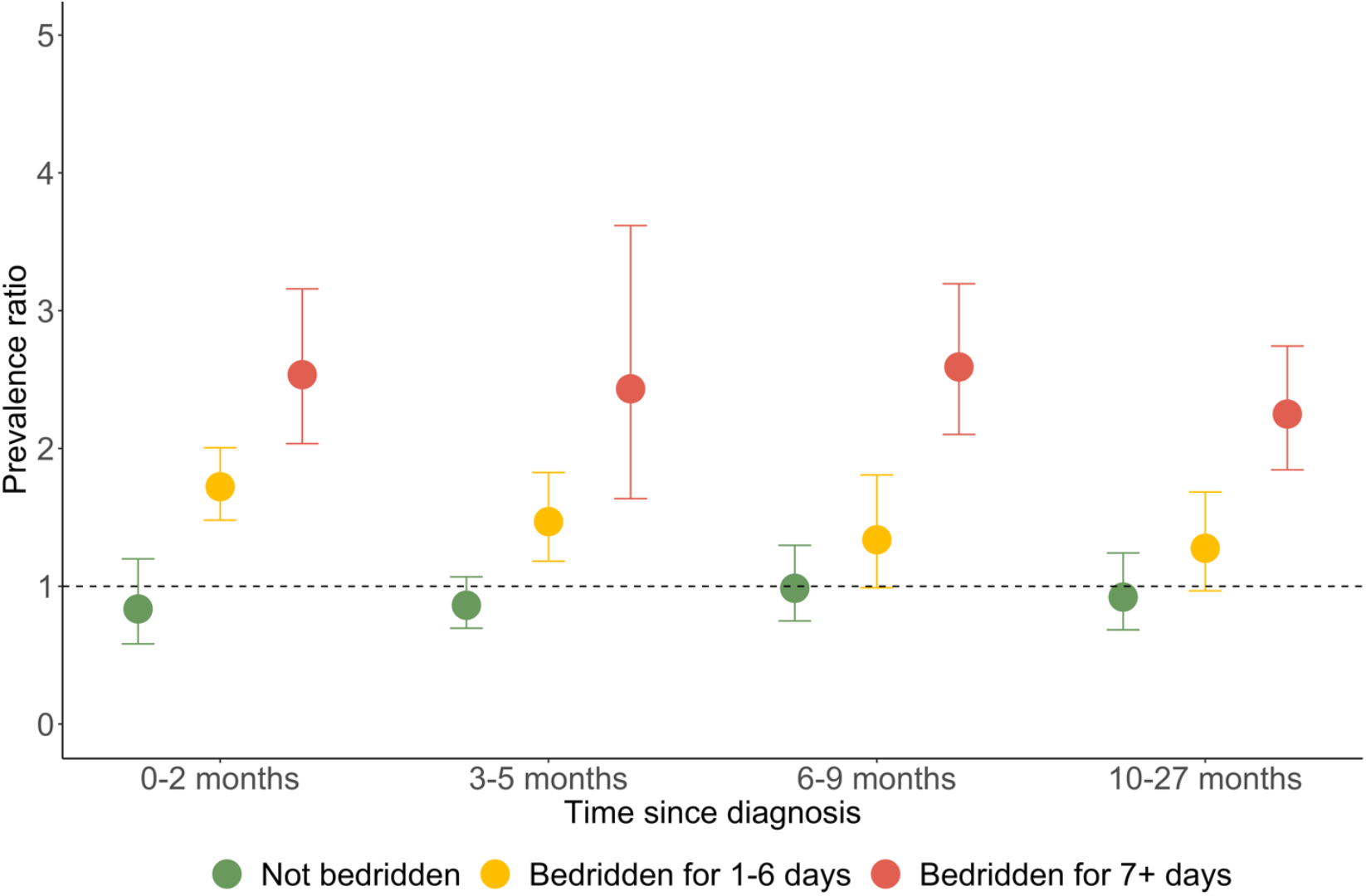
Prevalence ratio (95% confidence interval) of severe physical symptom burden (PHQ-15 ≥ 15) among people with COVID-19 compared with people *NOT* diagnosed with COVID-19, by illness severity (bedrriden) according to time from diagnosis, a meta-analysis combining four cohorts^a^. ^**a**^ Individuals with missing information on time since diagnosis or illness severity were excluded from this analysis (as shown in Table 1). Prevalence ratios were adjusted for age, gender, residency, average monthly income, current smoking, BMI, pre-existing comorbidity, relationship status, habitual drinking, previous diagnosis of psychiatric disorder, and response period. Income was not available in Omtanke2020 (SE), and relationship status was not available in DBDS (DK).

When investigating individual symptoms, we found the most prevalent symptoms to be headaches, low energy/fatigue, joint pain, trouble sleeping, and back pain (Supplementary Table S4). Compared with the non-COVID group, we found that a COVID-19 diagnosis was associated with a higher prevalence of several symptoms (Figure 3 A), including being bothered a lot due to shortness of breath (PR 2.15 [1.37-3.38]), dizziness (PR 1.58 [1.41-1.76]), heart racing (PR 1.55 [1.27-1.89]), headaches (PR 1.38 [1.23-1.54]), and back pain (PR 1.10 [1.05-1.17]) as well as being bothered a little due to chest pain (PR 1.34 [1.15-1.56]), low energy/fatigue (PR 1.08 [1.04-1.13]), and trouble sleeping (PR 1.04 [1.02-1.06]). Elevations in the prevalence of similar symptoms were noted in individual cohorts except for DBDS, where less symptoms were identified (Supplementary Figure S4). We observed a consistently higher prevalence, particularly for individuals bedridden 7 days or longer or hospitalized for COVID-19, among 8 of the 15 measured symptoms (Figure 4, Supplementary Table S5 and Figure S5). There was no clear decline in the prevalence increase over time for most symptoms, except for dizziness where the prevalence increase declined over time.

**Figure 3.**
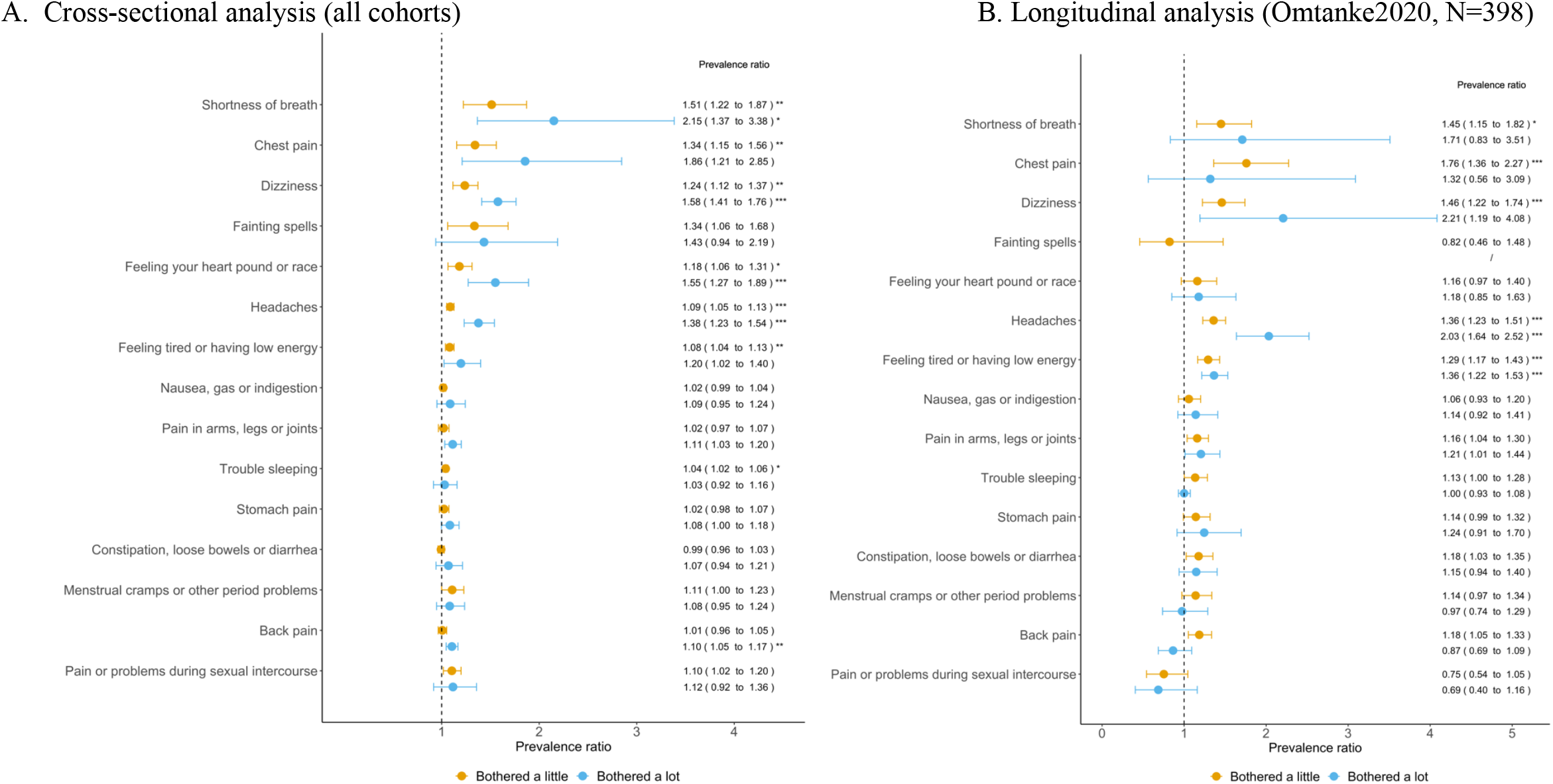
Prevalence ratio (95% confidence interval) of indivdual physical symptom severity among people with COVID-19 compared with those *NOT* diagnosed with COVID-19, by each physical symtom^a^. ^**a**^ Prevalence ratios were adjusted for age, gender, residency, average monthly income, current smoking, BMI, pre-existing comorbidity, relationship status, habitual drinking, previous diagnosis of psychiatric disorder, and response period. Income was not available in Omtanke2020 (SE), and relationship status was not available in DBDS (DK). Menstrual cramps were only applied to women aged <60 years. P-values were corrected for multiple testing using Bonferroni correction method. * indicates corrected P-value<0.05; **<0.01; ***<0.001.

**Figure 4.**
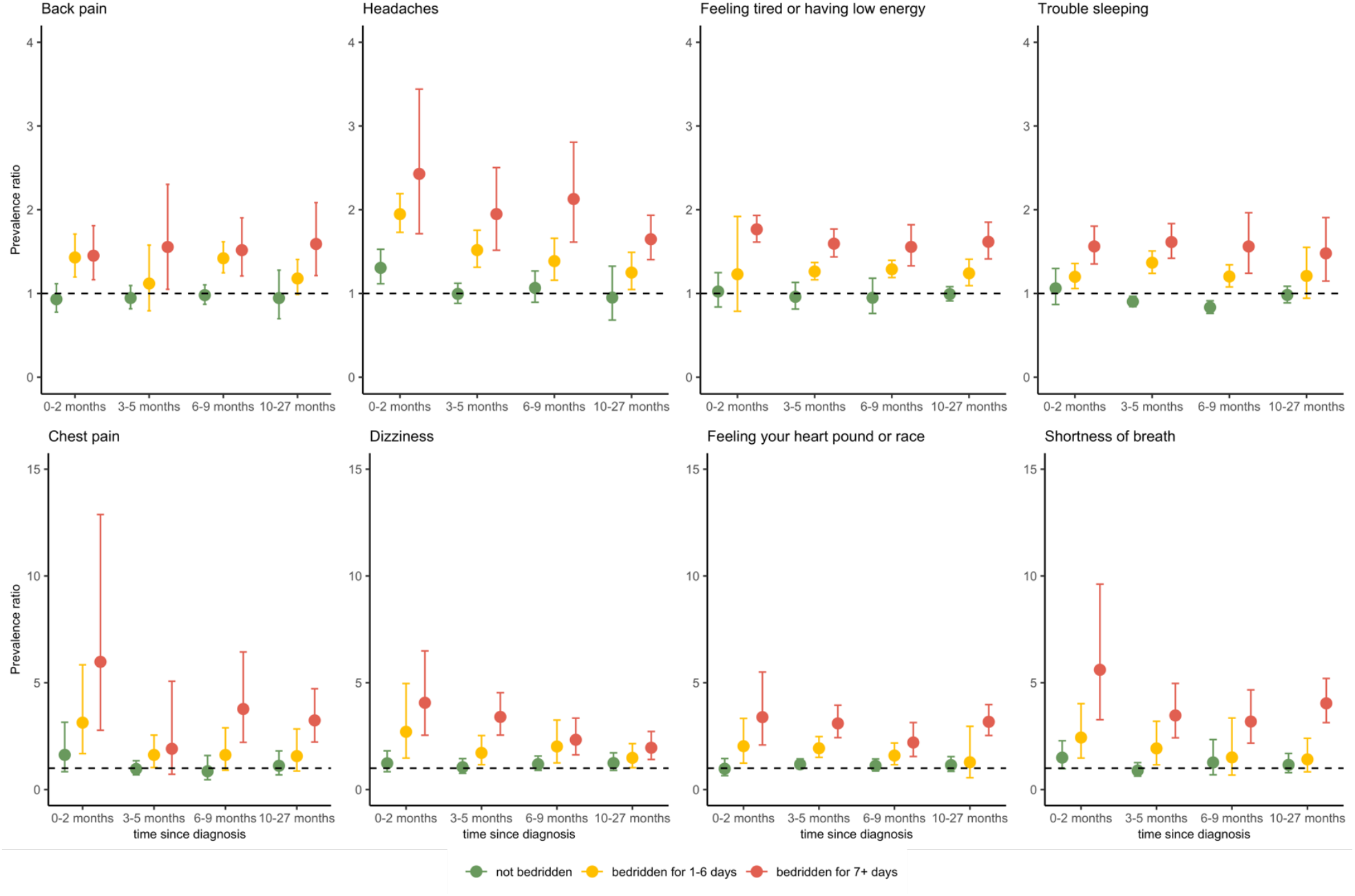
Prevalence ratio (95% confidence interval) of reporting *bothered a lot* to each symptom among people with COVID-19 compared with those *NOT* diagnosed with COVID-19 in C-19 Resilience, Omtanke2020 and DBDS, by illness severity (bedridden) according to time from diagnosis^a^. ^**a**^ Prevalence ratios were adjusted for age, gender, residency, average monthly income, current smoking, BMI, pre-existing comorbidity, relationship status, habitual drinking, previous diagnosis of psychiatric disorder, and response period. Income was not available in Omtanke2020 (SE), and relationship status was not available in DBDS (DK).

### Longitudinal analysis

The analysis of the subset of Omtanke2020 adult participants with pre-and post-COVID-19 measures of physical symptoms largely confirmed the results of our cross-sectional analysis. As compared to before diagnosis, we observed an elevation of being bothered a lot due to headaches (PR 2.03 [1.64-2.54]) and low energy/fatigue (PR 1.36 [1.22-1.53]) after diagnosis of COVID-19 (N=398, mean time interval=3.2 months) as well as being bothered a little due to shortness of breath (PR 1.45 [1.15-1.82]), chest pain (PR 1.76 [1.36-2.27]) and dizziness (PR 1.46 [1.22-1.74]) (Figure 3 B).

## Discussion

In this multinational observational study, we found an association between COVID-19 and persistence of severe physical symptoms up to 27 months following diagnosis. Overall, persons diagnosed with COVID-19 had a 37% higher prevalence of severe symptoms compared with those not diagnosed. The association was strongly modified by acute COVID-19 illness severity, as the prevalence increase was particularly great among individuals who were bedridden 7 days or more. The higher prevalence was observed for multiple symptoms, in particular shortness of breath, chest pain, dizziness, headaches, and low energy/fatigue. This finding was similarly identified in a longitudinal analysis comparing severe symptoms *before* and *after* diagnosis in Sweden. The persistently increased prevalence of multiple symptoms more than two years following severe COVID-19 demonstrates the importance of a sustained monitoring of physical symptoms in this patient group.

Long-term sequela following COVID-19 have been discussed continuously since the beginning of the pandemic. However, inconsistent definitions of post-COVID condition and a lack of comparison to individuals without COVID-19 diagnosis have made it difficult to comprehensively assess the long-term health impact of this infection. To our knowledge, this is the largest cohort study to quantify risks of common physical symptoms among individuals diagnosed with COVID-19 in the general population, and one of the few studies using a validated assessment tool. Our finding of increased risk of long-term severe symptoms is consistent with previous studies mostly based on hospitalized patients^5^ with a relatively short follow-up time of below^19,20^ or around ^21^ one year, and without a comparison to the population free of COVID-19.^4,19–21^ Consistent with our previous study,^10^ we utilized self-reported number of days bedridden in addition to hospitalization as a proxy for illness severity. As a result, we assessed severity of the acute phase of COVID-19 among non-hospitalized patients (92.4% of the COVID), which is usually not targeted in existing studies.^19^ Individuals with severe acute COVID-19, particularly those who were bedridden 7 days or longer (9.6% of the COVID group) or hospitalized (1.0% of the COVID group), had a greater increase in prevalence of severe symptoms, while those with a milder infection (not bedridden at all) showed none or only a marginal (bedridden 1-6 days) increase in such prevalence. The long-term health impact of severe COVID-19 is consistent with a recent hospital-based study from Italy, showing highly elevated prevalence in symptoms one year after acute illness.^21^ The ultimate duration of the risk increase remains unclear, and studies including long-term follow-ups are needed. Learning from the experiences from previous coronavirus outbreaks, similar persistence of physical symptoms have also been reported in survivors of SARS-CoV-1 (SARS), Middle East respiratory syndrome coronavirus (MERS), and acute respiratory distress syndrome (ARDS), including musculoskeletal pain, chronic fatigue, and exercise intolerance have previously been reported, up to 5 years after infection.^22–25^

Similar to previous studies,^4,19,26^ our findings indicate that shortness of breath, headaches, chest pain, dizziness, and low energy/fatigue may be core symptoms of Long COVID. The comparison on the magnitude of the increase in specific symptoms is however difficult across studies, due to the lack of comparison to individuals without COVID-19 in most of the previous studies.^4^ The findings on sustained increased prevalence of various symptoms after COVID-19, particularly after a severe course of acute illness, may be explained by several potential mechanisms. First, evidence shows that hyperactivity of proinflammatory cytokine response during the acute stage of the infection can cause damage to the musculoskeletal system, which can be associated with persistent symptoms such as fatigue, muscle weakness, and myalgias.^27^ Second, lingering cellular damages following acute infection can increase the levels of certain biomarkers including creatine kinase and thrombin, which regulate cellular energy metabolism and blood clotting, and can conceivably contribute to long-term physical symptoms observed following COVID-19.^27^ Third, chronic inflammation following acute infection can lead to alveolar damage in the lungs as well as microvascular injury.^27,28^ Also, there is suggestive evidence that some individuals do not completely clear the virus over time, resulting in persistent viral load in the body and chronic symptoms.^29^ In short, the prolonged symptoms following COVID-19 are likely to manifest multi-organ involvement,^30^ although these mechanisms are not fully understood. Further research is therefore needed to understand the long-term effects of the infection whereas sustained clinical surveillance on long-lasting physical symptoms is needed, by taking into account acute illness severity and time since diagnosis.

### Strengths and limitations

The strengths of our study include the large sample size with more than 22 000 persons diagnosed with COVID-19 with varying acute illness severity, as well as a large comparison group without COVID-19, across four Nordic countries. We were able to use detailed information on illness severity and time since diagnosis to investigate the association according to illness severity over time, with adjustment for a list of selected covariates. The validated questionnaire used to quantify the severity of physical symptoms enabled us to comprehensively assess the severe physical symptom burden, overall and individually. The more than 2-year follow-up provided, so far, the longest assessment on risk of persistent severe symptoms after infection with SARS-CoV-2. We were able to directly compare severe symptoms before and after COVID-19 diagnosis in a subset of Omtanke2020 participants, demonstrating a longitudinal impact of acute illness on participant’s physical health.

There are also some limitations to be noted. First, our study relies on self-reported information on COVID-19 diagnosis and physical symptoms, and therefore may suffer from recall bias, meaning that participants may not remember all the details of their illness. This will likely have diluted the results toward null. Second, individuals with COVID-19 may be more prone to report symptoms than the non-COVID group. Yet, the null association observed in the group who were diagnosed with COVID-19 but were not bedridden alleviates this concern to some extent. Third, the varying time periods of data collection in the four cohorts, as well as the varying waves of the pandemic across countries, resulted in substantial differences across cohorts and between COVID and non-COVID groups. We adjusted for the response period in the multivariable models to control for this discrepancy and the similar positive associations noted across the four cohorts argue against our findings being explained entirely by these factors. Fourth, despite consistent positive overall associations noted in each cohort, we found the prevalence increase of severe symptoms to be smaller in DBDS than C-19 Resilience (26% vs. 60% increase in prevalence). The DBDS cohort is comprised of active or former blood donors, presumably much healthier than the general population. This healthy donor effect may partly explain the smaller prevalence increase observed in DBDS, compared to the other cohorts. Apart from the abovementioned differences in the cohorts, the potential protective effect of the COVID-19 vaccine might also have played a role in participants with a late response period in Omtanke2020, MAP-19, and DBDS. Evidence has indeed shown reduced physical symptoms among fully vaccinated individuals.^21^ Future studies need to take into account the potential impact of vaccines and new SARS-CoV-2 variants on the risk of persistent physical symptoms. Lastly, our findings should be generalized with caution to other populations or countries with different societal, background, or healthcare systems than Nordic countries.

## Conclusion

This multinational study of persons with COVID-19 in four Nordic countries indicates that severe acute illness of SARS-CoV-2 infection is an important predictor of persistent physical symptoms, e.g., shortness of breath, chest pain, dizziness, headaches, and low energy/fatigue, up to 27 months after diagnosis. These findings highlight the importance of continued monitoring and alleviation of physical symptoms, at least during the first 2 years after diagnosis, among individuals suffering severe forms of COVID-19.

## Data Availability

The individual-level data underlying this article were subject to ethical approval and cannot be shared publicly due to data protection laws in each participating country.

## Author contributions

The participating COVIDMENT cohorts and/or their data collections were designed by QS, EEJ, OVE, MD, OBVP, SUJ, FF, UAV and their respective teams. UAV, QS and TA directed the combined effort of this study implementation. UAV, QS, EEJ and TA designed the analytical strategy in close collaboration with all team members and all authors helped to interpret the findings. QS, EEJ, OVE, MD conducted the literature review and drafted the manuscript under supervision of UAV. All authors revised the manuscript for critical content and approved the final version of the manuscript.

