## Supplementary file for "COVID-19 illness severity and 2-year prevalence of physical symptoms: an observational study in Iceland, Sweden, Norway and Denmark"

**Supplementary appendix**

**Supplementary table**

**Table S1 Overview of various measures and available variables used in each cohort.**

|  | **Iceland**  C-19 Resilience | **Sweden**  Omtanke2020 | **Norway**  MAP-19 | **Denmark**  DBDS |
| --- | --- | --- | --- | --- |
| **Ethical approval** |  |  |  |  |
|  | National Bioethics Committee (NBC no. 20–073, 21–071) as well as the National Data Protection Authority | Ethical approval no. 2020–01785 | Regional Committee for Medical Research Ethics, reference number: 125510 | Zealand and Central Denmark Regional Committees on Health Research Ethics (SJ-740 and M-2009237) and the Data Protection Agency (P-2019–99) |
| **COVID-19 related information** |  |  |  |  |
| COVID-19 diagnosis | Self-report of confirmed PCR-test | Self-report of confirmed PCR-test | Self-report of confirmed PCR-test | Self-report of confirmed PCR-test |
| COVID-19 illness severity - bedridden | Self-report of never bedridden/Bedridden 1-6 days/Bedridden 7 days or more | Self-report of never bedridden/Bedridden 1-6 days/Bedridden 7 days or more (asked at long surveys (baseline and 6-months follow-up) until June 2021, afterwards asked in all surveys) | Self-report of never bedridden/Bedridden 1-6 days/Bedridden 7 days or more | Self-report of never bedridden/Bedridden 1-6 days/Bedridden 7 days or more |
| COVID-19 illness severity - hospitalization | Self-report of not hospitalized/Hospitalized (non-ICU)/Hospitalized (ICU) | Self-report of not hospitalized/Hospitalized (non-ICU)/Hospitalized (ICU) | Self-report of not hospitalized/Hospitalized (no information on ICU) | Self-report of not hospitalized/Hospitalized (no information on ICU) |
| **Physical health instrument** |  |  |  |  |
| Severity of physical symptoms | Patient Health Questionnaire, 15-item (PHQ-15) | Patient Health Questionnaire, 15-item (PHQ-15) | Patient Health Questionnaire, 15-item (PHQ-15) | Patient Health Questionnaire, 15-item (PHQ-15) |
| Time for data collection | From April 2020 to August 2021 | From July 2021 to February 2022 | During March 2022 | From June to August 2022 |
| **Available covariates** |  |  |  |  |
| Age | Yes | Yes | Yes | Yes |
| Gender | Yes | Yes | Yes | Yes |
| Income | Yes | Not measured | Yes | Yes |
| Residence | Yes | Yes | Yes | Yes |
| Relationship status | Yes | Yes | Yes | Not measured |
| Current smoking | Yes | Yes | Yes | Yes |
| BMI | Yes | Yes | Yes | Yes |
| Habitual drinking | Yes | Yes | Yes | Yes |
| History of psychiatric disorder^b^ | Yes | Yes | Yes | Yes |
| Pre-existing comorbidity ^a^ | Yes | Yes | Yes | Yes |
| Current depressive symptoms | Yes | Yes | Yes | Yes |
| Current anxiety symptoms | Yes | Yes | Yes | Yes |
| Current COVID-19-related distress symptoms | Yes | Yes | Yes | Not measured |
| Response period | Yes | Yes | Yes | Yes |

^a^Pre-existing comorbidity was defined as hypertension, diabetes, heart disease, lung disease, chronic kidney disease, cancer, immunosuppressive state or immunosuppressive therapy, and was obtained from self-reported questionnaire in C-19 Resilience, Omtanke2020 and MAP-19. In DBDS, it was defined as having one or more of these comorbidities ascertained from the National Patient Register [ (astma (J45), diabetes (E10-E14), hypertension (I10-I16), heart disease (I20-I25), Stroke (I639), Osteoarthritis (M15-M19), Rheumatoid artritis (M059), Osteoporosis (M800, M810), Cancer (C chapter)].

^b^History of psychiatric disorder was obtained from self-reported questionnaire in C-19 Resilience, Omtanke2020 and MAP-19, and was ascertained from the National Patient Register (ICD-10 F chapter) and Prescription Register (ATC codes N05A, N06A, N05B, NA06B) in DBDS.

**Table S2. Background characteristics of study participants with and without a COVID-19 diagnosis in the four cohorts, separately and combined.**

|  | C-19 Resillience (IS) | | Omtanke2020 (SE) | | MAP-19 (NO) | | DBDS (DK) | | Overall | |
| --- | --- | --- | --- | --- | --- | --- | --- | --- | --- | --- |
|  | (N=14 358, median follow-up 133 days) | | (N=18 190, median follow-up 244 days) | | (N=3 310, median follow-up 26 days) | | (N=29 958, median follow-up 101 days) | | (N=64 880) | |
|  | non-COVID group (%) | COVID-19 group (%) | non-COVID group (%) | COVID-19 group (%) | non-COVID group (%) | COVID-19 group (%) | non-COVID group (%) | COVID-19 group (%) | non-COVID group (%) | COVID-19 group (%) |
| Total | 13284 (92.5%) | 1074 (7.5%) | 14 841 (81.6%) | 3 349 (18.4%) | 1 701 (51.4%) | 1 609 (48.6%) | 12 672 (43.3%) | 16 350 (56.7%) | 42 498 (65.5%) | 22 382 (34.5%) |
| Gender |  |  |  |  |  |  |  |  |  |  |
| Male | 3900 (29.4%) | 401 (37.3%) | 2734 (18.4%) | 594 (17.7%) | 363 (21.3%) | 310 (19.3%) | 6293 (49.7%) | 7202 (44.0%) | 13 290 (31.3%) | 8 507 (38.0%) |
| Female | 9307 (70.1%) | 663 (61.7%) | 12107 (81.6%) | 2755 (82.3%) | 1321 (77.7%) | 1290 (80.2%) | 6379 (50.3%) | 9148 (56.0%) | 29 114 (68.5%) | 13 856 (61.9%) |
| Other | 77 (0.6%) | 10 (0.9%) |  |  | 3 (0.2%) | 3 (0.2%) |  |  | 80 (0.2%) | 13 (0.1%) |
| Missing |  |  |  |  | 14 (0.8%) | 6 (0.4%) |  |  | 14 (0.0%) | 6 (0.0%) |
| Age |  |  |  |  |  |  |  |  |  |  |
| Mean (SD) | 55.4 (13.8) | 48.5 (14.8) | 52.0 (15.7) | 45.7 (13.2) | 38.9 (14.4) | 36.8 (13.4) | 56.6 (14.3) | 50.6 (14.3) | 54 (14.7) | 49 (11.7) |
| 18-29 years | 636 (4.8%) | 150 (14.0%) | 1409 (9.5%) | 427 (12.8%) | 586 (34.5%) | 623 (38.7%) | 703 (5.5%) | 1417 (8.7%) | 3 334 (7.8%) | 2 617 (11.7%) |
| 30-39 years | 1255 (9.4%) | 143 (13.3%) | 2316 (15.6%) | 736 (22.0%) | 403 (23.7%) | 377 (23.4%) | 1143 (9.0%) | 2515 (15.4%) | 5 117 (12.0%) | 3 771 (16.8%) |
| 40-49 years | 2303 (17.3%) | 240 (22.3%) | 2612 (17.6%) | 824 (24.6%) | 277 (16.3%) | 294 (18.3%) | 1584 (12.5%) | 3338 (20.4%) | 6 776 (15.9%) | 4 696 (21.0%) |
| 50-59 years | 3413 (25.7%) | 275 (25.6%) | 3242 (21.8%) | 847 (25.3%) | 223 (13.1%) | 193 (12.0%) | 3101 (24.5%) | 4195 (25.7%) | 9 979 (23.5%) | 5 510 (24.6%) |
| 60-69 years | 3611 (27.2%) | 190 (17.7%) | 2903 (19.6%) | 388 (11.6%) | 152 (8.9%) | 95 (5.9%) | 3643 (28.7%) | 3332 (20.4%) | 10 309 (24.3%) | 4 005 (17.9%) |
| 70 years or older | 2066 (15.6%) | 76 (7.1%) | 2359 (15.9%) | 127 (3.8%) | 46 (2.7%) | 20 (1.2%) | 2497 (19.7%) | 1553 (9.5%) | 6 968 (16.4%) | 1 776 (7.9%) |
| missing |  |  |  |  | 14 (0.8%) | 7 (0.4%) | <5 (0.0%) | <5 (0.0%) | 14 (<0.1%) | 7 (<0.1%) |
| Average monthly income |  |  |  |  |  |  |  |  |  |  |
| Low income | 2186 (16.5%) | 214 (19.9%) | - | - | 86 (5.1%) | 110 (6.8%) | 3227 (25.5%) | 2462 (15.1%) | 5 499 (12.9%) | 2 786 (12.4%) |
| Low-medium income | 3732 (28.1%) | 232 (21.6%) | - | - | 214 (12.6%) | 123 (7.6%) | 2315 (18.3%) | 3460 (21.2%) | 6 261 (14.7%) | 3 815 (17.0%) |
| Medium income | 3299 (24.8%) | 251 (23.4%) | - | - | 713 (20.2%) | 720 (19.8%) | 2350 (18.5%) | 3492 (21.4%) | 6 362 (15.0%) | 4 463 (19.9%) |
| Medium-high income | 2289 (17.2%) | 200 (18.6%) | - | - | - | - | 2442 (19.3%) | 3405 (20.8%) | 4 731 (11.1%) | 3 605 (16.1%) |
| High income | 1080 (8.1%) | 119 (11.1%) | - | - | 188 (11.1%) | 198 (12.3%) | 2338 (18.5%) | 3531 (21.6%) | 3 606 (8.5%) | 3 848 (17.2%) |
| Missing | 698 (5.3%) | 58 (5.4%) | - | - | 500 (29.4%) | 458 (28.5%) | - | - | 1 198 (2.8%) | 516 (2.3%) |
| Not measured | - | - | 14 841 (100.0) | 3 349 (100.0) | - | - | - | - | 14 841 (34.9%) | 3 349 (15.0%) |
| Residence |  |  |  |  |  |  |  |  |  |  |
| Capital area | 9065 (68.2%) | 784 (73.0%) | 6826 (46.0%) | 1744 (52.1%) | 456 (26.8%) | 475 (29.5%) | 5197 (41.0%) | 6749 (41.3%) | 21 544 (50.7%) | 9 752 (43.6%) |
| Elsewhere | 4088 (30.8%) | 279 (26.0%) | 7941 (53.5%) | 1583 (47.3%) | 1231 (72.4%) | 1126 (70.0%) | 7475 (59.0%) | 9601 (58.7%) | 20 735 (48.8%) | 12 589 (56.2%) |
| Missing or abroad | 131 (1.0%) | 11 (1.0%) | 74 (0.5%) | 22 (0.7%) | 14 (0.8%) | 8 (0.5%) | - | - | 219 (0.5%) | 41 (0.2%) |
| Relationship status |  |  |  |  |  |  |  |  |  |  |
| Single | 3224 (24.3%) | 229 (21.3%) | 4041 (27.2%) | 744 (22.2%) | 660 (38.8%) | 620 (38.5%) | - | - | 7 925 (18.6%) | 1 593 (7.1%) |
| In a relationship | 9964 (75.0%) | 833 (77.6%) | 10669 (71.9%) | 2566 (76.6%) | 1027 (60.4%) | 981 (61.0%) | - | - | 21 660 (51.0%) | 4 380 (19.6%) |
| Missing | 96 (0.7%) | 12 (1.1%) | 131 (0.9%) | 39 (1.2%) | 14 (0.8%) | 8 (0.5%) | - | - | 241 (0.6%) | 59 (0.3%) |
| Not measured | - | - | - | - | - | - | 12672 (100.0) | 16350 (100.0) | 12 672 (29.8%) | 16 350 (73.0%) |
| BMI (kg/m^2) |  |  |  |  |  |  |  |  |  |  |
| < 25, Normal or low weight | 3733 (28.1%) | 337 (31.4%) | 7787 (52.5%) | 1767 (52.8%) | 677 (39.8%) | 762 (47.4%) | 5394 (42.6%) | 7224 (44.2%) | 17 591 (41.4%) | 10 090 (45.1%) |
| 25-30, Overweight | 5115 (38.5%) | 380 (35.4%) | 4399 (29.6%) | 975 (29.1%) | 594 (34.9%) | 535 (33.3%) | 4796 (37.8%) | 6145 (37.6%) | 14 904 (35.1%) | 8 035 (35.9%) |
| > 30, Obese | 4080 (30.7%) | 298 (27.7%) | 1871 (12.6%) | 430 (12.8%) | 430 (25.3%) | 312 (19.4%) | 2327 (18.4%) | 2818 (17.2%) | 8 708 (20.5%) | 3 858 (17.2%) |
| Missing | 356 (2.7%) | 59 (5.5%) | 784 (5.3%) | 177 (5.3%) | - | - | 155 (1.2%) | 163 (1.0%) | 1 295 (3.0%) | 399 (1.8%) |
| Smoking status |  |  |  |  |  |  |  |  |  |  |
| No | 11510 (86.6%) | 979 (91.2%) | 12555 (84.6%) | 2740 (81.8%) | 1546 (90.9%) | 1527 (94.9%) | 11237 (88.7%) | 15033 (91.9%) | 36 848 (86.7%) | 20 279 (90.6%) |
| Yes | 1627 (12.2%) | 81 (7.5%) | 2019 (13.6%) | 533 (15.9%) | 155 (9.1%) | 82 (5.1%) | 1435 (11.3%) | 1317 (8.1%) | 5 236 (12.3%) | 2 013 (9.0%) |
| Missing | 147 (1.1%) | 14 (1.3%) | 267 (1.8%) | 76 (2.3%) | - | - | - | - | 414 (1.0%) | 90 (0.4%) |
| Habitual drinking |  |  |  |  |  |  |  |  |  |  |
| No | 11875 (89.4%) | 890 (82.9%) | 11099 (74.8%) | 2468 (73.7%) | 1666 (97.9%) | 1572 (97.7%) | 10815 (85.4%) | 13654 (83.5%) | 35 455 (83.4%) | 18 584 (83.0%) |
| Yes | 1409 (10.6%) | 184 (17.1%) | 3742 (25.2%) | 881 (26.3%) | 35 (2.1%) | 37 (2.3%) | 1857 (14.6%) | 2696 (16.5%) | 7 043 (16.6%) | 3 798 (17.0%) |
| History of psychiatric disorder^a^ |  |  |  |  |  |  |  |  |  |  |
| No | 9261 (69.7%) | 779 (72.5%) | 9803 (66.1%) | 2135 (63.8%) | 1249 (73.4%) | 1304 (81.0%) | 10873 (85.8%) | 14254 (87.2%) | 31 186 (73.4%) | 18 472 (82.5%) |
| Yes | 3798 (28.6%) | 275 (25.6%) | 4656 (31.4%) | 1114 (33.3%) | 452 (26.6%) | 305 (19.0%) | 1799 (14.2%) | 2096 (12.8%) | 10 705 (25.2%) | 3 790 (16.9%) |
| Missing | 225 (1.7%) | 20 (1.9%) | 382 (2.6%) | 100 (3.0%) |  |  |  |  | 607 (1.4%) | 120 (0.5%) |
| Pre-existing comorbidity^b^ |  |  |  |  |  |  |  |  |  |  |
| No | 9006 (67.8%) | 792 (73.7%) | 9725 (65.5%) | 2396 (71.5%) | 1048 (61.6%) | 1148 (71.3%) | 10248 (80.9%) | 13963 (85.4%) | 30 027 (70.7%) | 18 299 (81.8%) |
| Yes | 4278 (32.2%) | 282 (26.1%) | 4877 (32.9%) | 881 (26.4%) | 653 (38.4%) | 461 (28.7%) | 2348 (18.5%) | 2340 (14.3%) | 12 156 (28.6%) | 3 964 (17.7%) |
| Missing | - | - | 239 (1.6%) | 72 (2.1%) | - | - | 76 (0.6%) | 47 (0.3%) | 315 (0.7%) | 119 (0.5%) |
| Current depressive symptoms |  |  |  |  |  |  |  |  |  |  |
| No | 10668 (80.3%) | 817 (76.1%) | 12895 (86.9%) | 2793 (83.4%) | 1262 (74.2%) | 1209 (75.1%) | 11349 (89.6%) | 14578 (89.2%) | 36 174 (85.1%) | 19 397 (86.7%) |
| Yes | 2051 (15.4%) | 207 (19.3%) | 1933 (13.0%) | 554 (16.5%) | 439 (25.8%) | 400 (24.9%) | 1023 (8.1%) | 1483 (9.1%) | 5 446 (12.8%) | 2 644 (11.8%) |
| Missing | 565 (4.3%) | 50 (4.7%) | 13 (0.1%) | 2 (0.1%) | - | - | 300 (2.4%) | 289 (1.8%) | 878 (2.1%) | 341 (1.5%) |
| Current anxiety symptoms |  |  |  |  |  |  |  |  |  |  |
| No | 11528 (86.8%) | 931 (86.7%) | 13508 (91.0%) | 2977 (88.9%) | 1440 (84.7%) | 1402 (87.1%) | 12329 (97.3%) | 15836 (96.9%) | 38 805 (91.3%) | 21 146 (94.5%) |
| Yes | 1447 (10.9%) | 122 (11.4%) | 1309 (8.8%) | 366 (10.9%) | 261 (15.3%) | 207 (12.9%) | 343 (2.7%) | 514 (3.1%) | 3 360 (7.9%) | 1 209 (5.4%) |
| Missing | 309 (2.3%) | 21 (2.0%) | 24 (0.2%) | 6 (0.2%) | - | - | - | - | 333 (0.8%) | 27 (0.1%) |
| Current COVID-19-related distress symptoms |  |  |  |  |  |  |  |  |  |  |
| No | 8105 (61.0%) | 677 (63.0%) | 12018 (81.0%) | 2744 (81.9%) | 1408 (82.8%) | 1419 (88.2%) | - | - | 21 531 (50.7%) | 4 840 (21.6%) |
| Yes | 4861 (36.6%) | 375 (34.9%) | 2807 (18.9%) | 598 (17.9%) | 293 (17.2%) | 190 (11.8%) | - | - | 7 961 (18.7%) | 1 163 (5.2%) |
| Missing | 318 (2.4%) | 22 (2.0%) | 16 (0.1%) | 7 (0.2%) | - | - | - | - | 334 (0.8%) | 29 (0.1%) |
| Not measured | - | - | - | - | - | - | 12672 (100.0) | 16350 (100.0) | 12 672 (29.8%) | 16 350 (73.0%) |
| Response period |  |  |  |  |  |  |  |  |  |  |
| June 2020 or earlier | 12744 (95.9%) | 235 (21.9%) | - | - | - | - | - | - | 12 744 (30.0%) | 235 (1.0%) |
| July-September 2020 | 136 (1.0%) | 6 (0.6%) | - | - | - | - | - | - | 136 (0.3%) | 6 (0.0%) |
| October-December 2020 | 363 (2.7%) | 178 (16.6%) | - | - | - | - | - | - | 363 (0.9%) | 178 (0.8%) |
| January-March 2021 | 37 (0.3%) | 618 (57.5%) | - | - | - | - | - | - | 37 (0.1%) | 618 (2.8%) |
| April-June 2021 | 4 (0.0%) | 37 (3.4%) | - | - | - | - | - | - | 4 (0.0%) | 37 (0.2%) |
| July-September 2021 | - | - | 10072 (67.9%) | 1434 (42.8%) | - | - | - | - | 10 072 (23.7%) | 1 434 (6.4%) |
| October-December 2021 | - | - | 1970 (13.3%) | 423 (12.6%) | - | - | - | - | 1 970 (4.6%) | 423 (1.9%) |
| After January 2022 | - | - | 2799 (18.9%) | 1492 (44.6%) | 1 701 (100) | 1 609 (100) | 12672 (100.0) | 16350 (100.0) | 17 172 (40.4%) | 19 451 (86.9%) |
| Bedridden |  |  |  |  |  |  |  |  |  |  |
| Not bedridden | - | 417 (38.8%) | - | 901 (26.9%) | - | 559 (34.7%) | - | 13150 (80.4%) | - | 15 027 (67.1%) |
| Bedridden 1-6 days | - | 424 (39.5%) | - | 965 (28.8%) | - | 939 (58.4%) | - | 1799 (11.0%) | - | 4 127 (18.4%) |
| Bedridden 7 days or more | - | 233 (21.7%) | - | 403 (12.0%) | - | 111 (6.9%) | - | 1401 (8.6%) | - | 2 148 (9.6%) |
| missing | - |  | - | 1080 (32.2%) | - |  | - | - | - | 1 080 (4.8%) |
| Hospitalization |  |  |  |  |  |  |  |  |  |  |
| Not hospitalized | - | 577 (53.7%) | - | 2336 (69.8%) | - | 1479 (91.9%) | - | 16278 (99.6%) | - | 20670 (92,4%) |
| Hospitalized | - | 80 (7.4%) | - | 75 (2.2%) | - | 11 (0.7%) | - | 64 (0.4%) | - | 230 (1,0%) |
| Missing | - | 417 (38.8%) | - | 938 (28.0%) | - | 119 (7.4%) | - | 8 (0.0%) | - | 1482 (6,6%) |
| Time since COVID-19 diagnosis |  |  |  |  |  |  |  |  |  |  |
| 0-2 months | - | 185 (17.2%) | - | 993 (29.7%) | - | 1154 (71.7%) | - | 818 (5.0%) | - | 3 150 (14.1%) |
| 3-5 months | - | 657 (61.2%) | - | 614 (18.3%) | - | 335 (20.8%) | - | 9273 (56.7%) | - | 10 879 (48.6%) |
| 6-9 months | - | 89 (8.3%) | - | 744 (22.2%) | - | 27 (1.7%) | - | 3962 (24.2%) | - | 4 822 (21.5%) |
| 10-27 months | - | 137 (12.8%) | - | 715 (21.3%) | - | 93 (5.8%) | - | 2297 (14.0%) | - | 3 242 (14.5%) |
| Missing | - | 6 (0.6%) | - | 283 (8.5%) | - |  | - | - | - | 289 (1.3%) |

^a^History of psychiatric disorder was obtained from self-reported questionnaire in C-19 Resilience, Omtanke2020 and MAP-19, and was ascertained from the National Patient Register (ICD-10 F chapter) and Prescription Register (ATC codes N05A, N06A, N05B, NA06B) in DBDS.

^b^ Pre-existing comorbidity was defined as hypertension, diabetes, heart disease, lung disease, chronic kidney disease, cancer, immunosuppressive state or immunosuppressive therapy, and was obtained from self-reported questionnaire in C-19 Resilience, Omtanke2020 and MAP-19. In DBDS, it was defined as having one or more of these comorbidities ascertained from the National Patient Register [ (astma (J45), diabetes (E10-E14), hypertension (I10-I16), heart disease (I20-I25), Stroke (I639), Osteoarthritis (M15-M19), Rheumatoid artritis (M059), Osteoporosis (M800, M810), Cancer (C chapter)].

**Table S3. Prevalence of severe physical symptom burden (PHQ-15≥15) among individuals with and without COVID-19 in the four cohorts, according to gender, age, current mental health indicators (current depressive, anxiety, and COVID-19 related distress symtoms) and pre-existing comorbidity, and by time since diagnosis and illness severity among those with COVID-19.**

|  | C-19 Resillience (IS) | | Omtanke2020 (SE) | | MAP-19 (NO) | | DBDS (DK) | |
| --- | --- | --- | --- | --- | --- | --- | --- | --- |
|  | non-COVID group (%) | COVID-19 group (%) | non-COVID group (%) | COVID-19 group (%) | non-COVID group (%) | COVID-19 group (%) | non-COVID group (%) | COVID-19 group (%) |
| Total | 2080 (9.7%) | 251 (16.0%) | 1346 (5.5%) | 380 (8.0%) | 129 (7.6%) | 137 (8.5%) | 154 (1.2%) | 267 (1.6%) |
| Gender |  |  |  |  |  |  |  |  |
| Male | 269 (4.2%) | 46 (7.8%) | 93 (2.1%) | 30 (3.5%) | 22 (6.1%) | 23 (7.4%) | 46 (0.7%) | 47 (0.7%) |
| Female | 1792 (12.0%) | 202 (21.0%) | 1253 (6.2%) | 350 (9.0%) | 107 (8.1%) | 114 (8.8%) | 108 (1.7%) | 220 (2.4%) |
| Age |  |  |  |  |  |  |  |  |
| 18-39 years | 496 (17.2%) | 84 (21.9%) | 473 (8.0%) | 175 (11.3%) | 76 (7.7%) | 100 (10.0%) | 48 (2.6%) | 105 (2.7%) |
| 40-59 years | 974 (10.7%) | 124 (16.2%) | 590 (6.2%) | 167 (7.0%) | 39 (7.8%) | 31 (6.4%) | 60 (1.3%) | 115 (1.5%) |
| 60 years or older | 610 (6.5%) | 43 (10.3%) | 283 (3.1%) | 38 (4.8%) | 14 (7.1%) | 6 (5.2%) | 46 (0.7%) | 47 (1.0%) |
| Current depressive symptoms |  |  |  |  |  |  |  |  |
| No | 707 (4.1%) | 83 (6.9%) | 530 (2.4%) | 163 (4.1%) | 23 (1.8%) | 34 (2.8%) | 42 (0.4%) | 77 (0.5%) |
| Yes | 1203 (38.0%) | 147 (50.5%) | 815 (27.4%) | 216 (29.7%) | 106 (24.1%) | 103 (25.8%) | 111 (10.9%) | 185 (12.5%) |
| Current anxiety symptoms |  |  |  |  |  |  |  |  |
| No | 1139 (6.0%) | 154 (11.3%) | 752 (3.3%) | 225 (5.3%) | 51 (3.5%) | 80 (5.7%) | 127 (1.0%) | 215 (1.4%) |
| Yes | 868 (40.5%) | 92 (55.1%) | 592 (29.4%) | 154 (32.2%) | 78 (29.9%) | 57 (27.5%) | 27 (7.9%) | 52 (10.1%) |
| Current COVID-19-related distress symptoms |  |  |  |  |  |  |  |  |
| No | 601 (4.2%) | 75 (7.2%) | 653 (3.3%) | 200 (5.1%) | 50 (3.6%) | 70 (4.9%) | - | - |
| Yes | 1409 (21.1%) | 175 (35.5%) | 688 (14.7%) | 178 (21.4%) | 79 (27.0%) | 67 (35.3%) | - | - |
| Pre-existing comorbidity |  |  |  |  |  |  |  |  |
| No | 1346 (9.3%) | 183 (16.0%) | 700 (4.3%) | 226 (6.7%) | 46 (4.4%) | 64 (5.6%) | 110 (1.1%) | 213 (1.5%) |
| Yes | 734 (10.5%) | 68 (16.1%) | 621 (7.6%) | 145 (11.3%) | 83 (12.7%) | 73 (15.8%) | 44 (1.8%) | 54 (2.3%) |
| By time since diagnosis according to illness severity (time bedridden) | | | | | | | | |
| 0-2 months |  |  |  |  |  |  |  |  |
| *Not bedridden* | - | 6 (8.7%) | - | 28 (6.6%) | - | 14 (3.4%) | - | 7 (1.2%) |
| *Bedridden 1-6 days* | - | 17 (24.6%) | - | 59 (12.1%) | - | 68 (9.9%) | - | <5 ^a^ |
| *Bedridden 7 days or more* | - | 13 (26.5%) | - | 16 (22.2%) | - | 22 (37.3%) | - | <5 ^a^ |
| 3-5 months |  |  |  |  |  |  |  |  |
| *Not bedridden* | - | 25 (9.7%) | - | 5 (4.4%) | - | 0 (0%) | - | 83 (1.1%) |
| *Bedridden 1-6 days* | - | 53 (19.3%) | - | 8 (7.8%) | - | 4 (8.0%) | - | 26 (2.6%) |
| *Bedridden 7 days or more* | - | 39 (28.3%) | - | 7 (12.1%) | - | 5 (20.0%) | - | 37 (7.0%) |
| 6-9 months |  |  |  |  |  |  |  |  |
| *Not bedridden* | - | 15 (11.2%) | - | 11 (5.6%) | - | 2 (1.6%) | - | 34 (1.1%) |
| *Bedridden 1-6 days* | - | 24 (15.6%) | - | 13 (6.4%) | - | 14 (7.4%) | - | 6 (1.3%) |
| *Bedridden 7 days or more* | - | 14 (26.9%) | - | 25 (20.3%) | - | 6 (26.1%) | - | 19 (5.9%) |
| 10-27 months |  |  |  |  |  |  |  |  |
| *Not bedridden* | - | 6 (4.3%) | - | 20 (5.5%) | - | 1 (9.1%) | - | 23 (1.4%) |
| *Bedridden 1-6 days* | - | 20 (17.9%) | - | 26 (8.0%) | - | 0 (0%) | - | <5 ^a^ |
| *Bedridden 7 days or more* | - | 17 (15.5%) | - | 49 (17.2%) | - | 1 (25.0%) | - | 23 (4.6%) |

^a^ Prevalence for number less than 5 was not reported in DBDS due to the data regulation in Denmark.

**Table S4. Prevalence of indivdual physical symptom severity among people with and without COVID-19 in four cohorts.**

|  | C-19 Resillience (IS) | Omtanke2020 (SE) | MAP-19 (NO) | DBDS (DK) |
| --- | --- | --- | --- | --- |
|  | non-COVID group (%) / COVID-19 group (%) | non-COVID group (%) / COVID-19 group (%) | non-COVID group (%) / COVID-19 group (%) | non-COVID group (%) / COVID-19 group (%) |
| Shortness of breath |  |  |  |  |
| Bothered a little | 3265 (15.2%) / 401 (25.6%) | 3232 (13.1%) / 954 (20.1%) | 431 (25.3%) / 629 (39.1%) | 1353 (10.7%) / 1695 (10.4%) |
| Bothered a lot | 518 (2.4%) / 89 (5.7%) | 414 (1.7%) / 155 (3.3%) | 62 (3.6%) / 138 (8.6%) | 173 (1.4%) / 206 (1.3%) |
| Chest pain |  |  |  |  |
| Bothered a little | 3044 (14.2%) / 316 (20.2%) | 2642 (10.7%) / 697 (14.7%) | 231 (13.6%) / 292 (18.1%) | 905 (7.1%) / 1271 (7.8%) |
| Bothered a lot | 325 (1.5%) / 55 (3.5%) | 233 (0.9%) / 95 (2.0%) | 17 (1.0%) / 37 (2.3%) | 8 (0.6%) / 110 (0.7%) |
| Dizziness |  |  |  |  |
| Bothered a little | 5545 (25.9%) / 508 (32.5%) | 5710 (23.2%) / 1278 (27.0%) | 481 (28.3%) / 587 (36.5%) | 1953 (15.4%) / 2711 (16.6%) |
| Bothered a lot | 854 (4.0%) / 106 (6.8%) | 629 (2.6%) / 186 (3.9%) | 70 (4.1%) / 86 (5.3%) | 160 (1.3%) / 272 (1.7%) |
| Fainting spells |  |  |  |  |
| Bothered a little | 370 (1.7%) / 42 (2.7%) | 659 (2.7%) / 194 (4.1%) | 43 (2.5%) / 44 (2.7%) | 101 (0.8%) / 123 (0.8%) |
| Bothered a lot | 41 (0.2%) / 2 (0.1%) | 58 (0.2%) / 19 (0.4%) | 0 (0%) / 5 (0.3%) | 15 (0.1%) / 20 (0.1%) |
| Feeling your heart pound or race |  |  |  |  |
| Bothered a little | 6549 (30.6%) / 571 (36.5%) | 4825 (19.6%) / 1149 (24.2%) | 361 (21.2%) / 361 (22.4%) | 2046 (16.1%) / 2750 (16.8%) |
| Bothered a lot | 1077 (5.0%) / 164 (10.5%) | 646 (2.6%) / 183 (3.9%) | 54 (3.2%) / 60 (3.7%) | 191 (1.5%) / 357 (2.2%) |
| Headaches |  |  |  |  |
| Bothered a little | 9009 (42.1%) / 693 (44.3%) | 11145 (45.2%) / 2416 (51.0%) | 831 (48.9%) / 848 (52.7%) | 3701 (29.2%) / 5738 (35.1%) |
| Bothered a lot | 2168 (10.1%) / 259 (16.5%) | 2169 (8.8%) / 665 (14.0%) | 194 (11.4%) / 263 (16.3%) | 447 (3.5%) / 846 (5.2%) |
| Feeling tired or having low energy |  |  |  |  |
| Bothered a little | 9565 (44.7%) / 623 (39.8%) | 11086 (45.0%) / 2118 (44.7%) | 783 (46.0%) / 766 (47.6%) | 3208 (25.3%) / 4653 (28.5%) |
| Bothered a lot | 5281 (24.7%) / 594 (38.0%) | 4801 (19.5%) / 1322 (27.9%) | 484 (28.5%) / 548 (34.1%) | 1504 (11.9%) / 2040 (12.5%) |
| Nausea, gas or indigestion |  |  |  |  |
| Bothered a little | 8654 (40.4%) / 641 (41.0%) | 8780 (35.6%) / 1750 (36.9%) | 731 (43.0%) / 710 (44.1%) | 2901 (22.9%) / 3752 (22.9%) |
| Bothered a lot | 2232 (10.4%) / 215 (13.7%) | 1960 (8.0%) / 471 (9.9%) | 150 (8.8%) / 130 (8.1%) | 416 (3.3%) / 568 (3.5%) |
| Pain in arms, legs or joints |  |  |  |  |
| Bothered a little | 9392 (43.8%) / 643 (41.1%) | 9637 (39.1%) / 1857 (39.2%) | 744 (43.7%) / 653 (40.6%) | 4875 (38.5%) / 5714 (34.9%) |
| Bothered a lot | 5592 (26.1%) / 447 (28.6%) | 3340 (13.5%) / 674 (14.2%) | 217 (12.8%) / 169 (10.5%) | 1387 (10.9%) / 1685 (10.3%) |
| Trouble sleeping |  |  |  |  |
| Bothered a little | 9334 (43.6%) / 693 (44.3%) | 10711 (43.4%) / 2147 (45.3%) | 686 (40.3%) / 658 (40.9%) | 4663 (36.8%) / 6863 (42.0%) |
| Bothered a lot | 4727 (22.1%) / 388 (24.8%) | 3482 (14.1%) / 783 (16.5%) | 318 (18.7%) / 222 (13.8%) | 1580 (12.5%) / 2438 (14.9%) |
| Stomach pain |  |  |  |  |
| Bothered a little | 7506 (35.0%) / 584 (37.3%) | 7551 (30.6%) / 1586 (33.5%) | 558 (32.8%) / 577 (35.9%) | 1574 (12.4%) / 2220 (13.6%) |
| Bothered a lot | 1680 (7.8%) / 148 (9.5%) | 1470 (6.0%) / 358 (7.6%) | 90 (5.3%) / 71 (4.4%) | 188 (1.5%) / 287 (1.8%) |
| Constipation, loose bowels or diarrhea |  |  |  |  |
| Bothered a little | 9095 (42.5%) / 674 (43.1%) | 8971 (36.4%) / 1757 (37.1%) | 745 (43.8%) / 699 (43.4%) | 2844 (22.4%) / 3701 (22.6%) |
| Bothered a lot | 2805 (13.1%) / 244 (15.6%) | 2069 (8.4%) / 473 (10.0%) | 163 (9.6%) / 134 (8.3%) | 439 (3.5%) / 592 (3.6%) |
| Menstrual cramps or other period problems |  |  |  |  |
| Bothered a little | 1924 (9.0%) / 200 (12.8%) | 3699 (28.4%) / 1006 (30.3%) | 299 (17.6%) / 373 (23.2%) | 752 (5.9%) / 1583 (9.7%) |
| Bothered a lot | 832 (3.9%) / 89 (5.7%) | 1268 (9.8%) / 324 (9.8%) | 85 (5.0%) / 93 (5.8%) | 202 (1.6%) / 441 (2.7%) |
| Back pain |  |  |  |  |
| Bothered a little | 8952 (41.8%) / 624 (39.9%) | 9571 (38.8%) / 1985 (41.9%) | 725 (42.6%) / 636 (39.5%) | 4201 (33.2%) / 5149 (31.5%) |
| Bothered a lot | 4044 (18.9%) / 343 (21.9%) | 2712 (11.0%) / 589 (12.4%) | 169 (9.9%) / 132 (8.2%) | 877 (6.9%) / 1198 (7.3%) |
| Pain or problems during sexual intercourse |  |  |  |  |
| Bothered a little | 1655 (7.7%) / 140 (8.9%) | 2819 (11.4%) / 563 (11.9%) | 102 (6.0%) / 107 (6.7%) | 653 (5.2%) / 1032 (6.3%) |
| Bothered a lot | 779 (3.6%) / 65 (4.2%) | 457 (1.9%) / 95 (2.0%) | 32 (1.9%) / 30 (1.9%) | 142 (1.1%) / 232 (1.4%) |

**Table S5. Prevalence of reporting *bothered a lot* to individual symptoms among people with and without COVID-19 in C-19 Resilience, Omtanke2020, and DBDS, by illness severity (bedridden) according to time from diagnosis^a^**

|  | C-19 Resillience (IS) | Omtanke2020 (SE) | DBDS (DK) |
| --- | --- | --- | --- |
| Back pain - Bothered a lot |  |  |  |
| non-COVID group (%) | 4044 (18.9%) | 2712 (11.0%) | 877 (6.9%) |
| COVID-19 group (%) | 343 (21.9%) | 589 (12.4%) | 1198 (7.3%) |
| 0-2 months |  |  |  |
| *Not bedridden* | 8 (11.6%) | 43 (10.1%) | 42 (6.9%) |
| *Bedridden 1-6 days* | 19 (27.5%) | 80 (16.4%) | 14 (8.9%) |
| *Bedridden 7 days or more* | 17 (34.7%) | 14 (19.4%) | <5 |
| 3-5 months |  |  |  |
| *Not bedridden* | 46 (17.9%) | 12 (10.6%) | 496 (6.4%) |
| *Bedridden 1-6 days* | 62 (22.6%) | 6 (5.9%) | 88 (8.8%) |
| *Bedridden 7 days or more* | 38 (27.5%) | 10 (17.2%) | 80 (15.2%) |
| 6-9 months |  |  |  |
| *Not bedridden* | 27 (20.1%) | 18 (9.1%) | 185 (5.8%) |
| *Bedridden 1-6 days* | 43 (27.9%) | 28 (13.7%) | 41 (9.2%) |
| *Bedridden 7 days or more* | 18 (34.6%) | 23 (18.7%) | 43 (13.4%) |
| 10-27 months |  |  |  |
| *Not bedridden* | 12 (8.6%) | 38 (10.4%) | 117 (7.3%) |
| *Bedridden 1-6 days* | 24 (21.4%) | 48 (14.7%) | 17 (8.7%) |
| *Bedridden 7 days or more* | 27 (24.5%) | 51 (17.9%) | 71 (14.2%) |
| Headaches - Bothered a lot |  |  |  |
| non-COVID group (%) | 2168 (10.1%) | 2169 (8.8%) | 447 (3.5%) |
| COVID-19 group (%) | 259 (16.5%) | 665 (14.0%) | 846 (5.2%) |
| 0-2 months |  |  |  |
| *Not bedridden* | 8 (11.6%) | 66 (15.5%) | 24 (4.0%) |
| *Bedridden 1-6 days* | 16 (23.2%) | 136 (27.8%) | 14 (8.9%) |
| *Bedridden 7 days or more* | 14 (28.6%) | 23 (31.9%) | 11 (19.6%) |
| 3-5 months |  |  |  |
| *Not bedridden* | 36 (14.0%) | 14 (12.4%) | 305 (3.9%) |
| *Bedridden 1-6 days* | 64 (23.4%) | 15 (14.7%) | 86 (8.6%) |
| *Bedridden 7 days or more* | 36 (26.1%) | 8 (13.8%) | 70 (13.3%) |
| 6-9 months |  |  |  |
| *Not bedridden* | 19 (14.2%) | 22 (11.2%) | 126 (3.9%) |
| *Bedridden 1-6 days* | 16 (10.4%) | 26 (12.7%) | 38 (8.5%) |
| *Bedridden 7 days or more* | 10 (19.2%) | 30 (24.4%) | 50 (15.6%) |
| 10-27 months |  |  |  |
| *Not bedridden* | 5 (3.6%) | 40 (11.0%) | 73 (4.6%) |
| *Bedridden 1-6 days* | 18 (16.1%) | 39 (12.0%) | 12 (6.1%) |
| *Bedridden 7 days or more* | 14 (12.7%) | 53 (18.6%) | 37 (7.4%) |
| Feeling tired or having low energy - Bothered a lot |  |  |  |
| non-COVID group (%) | 5281 (24.7%) | 4801 (19.5%) | 1504 (11.9%) |
| COVID-19 group (%) | 594 (38.0%) | 1322 (27.9%) | 2040 (12.5%) |
| 0-2 months |  |  |  |
| *Not bedridden* | 17 (24.6%) | 108 (25.4%) | 60 (9.9%) |
| *Bedridden 1-6 days* | 32 (46.4%) | 208 (42.5%) | 13 (8.3%) |
| *Bedridden 7 days or more* | 25 (51.0%) | 50 (69.4%) | 13 (23.2%) |
| 3-5 months |  |  |  |
| *Not bedridden* | 68 (26.5%) | 20 (17.7%) | 812 (10.5%) |
| *Bedridden 1-6 days* | 126 (46.0%) | 29 (28.4%) | 172 (17.2%) |
| *Bedridden 7 days or more* | 73 (52.9%) | 20 (34.5%) | 123 (23.4%) |
| 6-9 months |  |  |  |
| *Not bedridden* | 43 (32.1%) | 35 (17.8%) | 338 (10.6%) |
| *Bedridden 1-6 days* | 57 (37.0%) | 51 (25.0%) | 84 (18.8%) |
| *Bedridden 7 days or more* | 27 (51.9%) | 48 (39.0%) | 76 (23.8%) |
| 10-27 months |  |  |  |
| *Not bedridden* | 32 (23.0%) | 97 (26.6%) | 188 (11.7%) |
| *Bedridden 1-6 days* | 39 (34.8%) | 87 (26.7%) | 37 (18.9%) |
| *Bedridden 7 days or more* | 53 (48.2%) | 114 (40.0%) | 124 (24.8%) |
| Trouble sleeping - Bothered a lot |  |  |  |
| non-COVID group (%) | 4727 (22.1%) | 3482 (14.1%) | 1580 (12.5%) |
| COVID-19 group (%) | 388 (24.8%) | 783 (16.5%) | 2438 (14.9%) |
| 0-2 months |  |  |  |
| *Not bedridden* | 15 (21.7%) | 63 (14.8%) | 85 (14.0%) |
| *Bedridden 1-6 days* | 18 (26.1%) | 102 (20.9%) | 32 (20.4%) |
| *Bedridden 7 days or more* | 20 (40.8%) | 22 (30.6%) | 16 (28.6%) |
| 3-5 months |  |  |  |
| *Not bedridden* | 47 (18.3%) | 16 (14.2%) | 933 (12.0%) |
| *Bedridden 1-6 days* | 92 (33.6%) | 15 (14.7%) | 224 (22.4%) |
| *Bedridden 7 days or more* | 46 (33.3%) | 19 (32.8%) | 160 (30.4%) |
| 6-9 months |  |  |  |
| *Not bedridden* | 23 (17.2%) | 15 (7.6%) | 371 (11.6%) |
| *Bedridden 1-6 days* | 35 (22.7%) | 28 (13.7%) | 93 (20.9%) |
| *Bedridden 7 days or more* | 17 (32.7%) | 33 (26.8%) | 106 (33.1%) |
| 10-27 months |  |  |  |
| *Not bedridden* | 25 (18.0%) | 52 (14.2%) | 224 (14.0%) |
| *Bedridden 1-6 days* | 28 (25.0%) | 49 (15.0%) | 48 (24.5%) |
| *Bedridden 7 days or more* | 22 (20.0%) | 84 (29.5%) | 146 (29.3%) |
| Chest pain - Bothered a lot |  |  |  |
| non-COVID group (%) | 325 (1.5%) | 233 (0.9%) | 78 (0.6%) |
| COVID-19 group (%) | 55 (3.5%) | 95 (2.0%) | 110 (0.7%) |
| 0-2 months |  |  |  |
| *Not bedridden* | 2 (2.9%) | 7 (1.6%) | <5 |
| *Bedridden 1-6 days* | 6 (8.7%) | 12 (2.5%) | <5 |
| *Bedridden 7 days or more* | 4 (8.2%) | 7 (9.7%) | <5 |
| 3-5 months |  |  |  |
| *Not bedridden* | 6 (2.3%) | 2 (1.8%) | 42 (0.5%) |
| *Bedridden 1-6 days* | 11 (4.0%) | 2 (2.0%) | 9 (0.9%) |
| *Bedridden 7 days or more* | 9 (6.5%) | 1 (1.7%) | <5 |
| 6-9 months |  |  |  |
| *Not bedridden* | 3 (2.2%) | 2 (1.0%) | 12 (0.4%) |
| *Bedridden 1-6 days* | 5 (3.2%) | 2 (1.0%) | <5 |
| *Bedridden 7 days or more* | 0 | 4 (3.3%) | 10 (3.1%) |
| 10-27 months |  |  |  |
| *Not bedridden* | 1 (0.7%) | 4 (1.1%) | 13 (0.8%) |
| *Bedridden 1-6 days* | 3 (2.7%) | 5 (1.5%) | <5 |
| *Bedridden 7 days or more* | 5 (4.5%) | 12 (4.2%) | 11 (2.2%) |
| Dizziness - Bothered a lot |  |  |  |
| non-COVID group (%) | 854 (4.0%) | 629 (2.6%) | 160 (1.3%) |
| COVID-19 group (%) | 106 (6.8%) | 186 (3.9%) | 272 (1.7%) |
| 0-2 months |  |  |  |
| *Not bedridden* | 3 (4.3%) | 15 (3.5%) | 6 (1.0%) |
| *Bedridden 1-6 days* | 12 (17.4%) | 24 (4.9%) | <5 |
| *Bedridden 7 days or more* | 4 (8.2%) | 9 (12.5%) | 5 (8.9%) |
| 3-5 months |  |  |  |
| *Not bedridden* | 8 (3.1%) | 2 (1.8%) | 107 (1.4%) |
| *Bedridden 1-6 days* | 22 (8.0%) | 2 (2.0%) | 19 (1.9%) |
| *Bedridden 7 days or more* | 21 (15.2%) | 5 (8.6%) | 24 (4.6%) |
| 6-9 months |  |  |  |
| *Not bedridden* | 5 (3.7%) | 7 (3.6%) | 41 (1.3%) |
| *Bedridden 1-6 days* | 8 (5.2%) | 13 (6.4%) | 9 (2.0%) |
| *Bedridden 7 days or more* | 4 (7.7%) | 10 (8.1%) | 9 (2.8%) |
| 10-27 months |  |  |  |
| *Not bedridden* | 4 (2.9%) | 8 (2.2%) | 27 (1.7%) |
| *Bedridden 1-6 days* | 7 (6.3%) | 12 (3.7%) | <5 |
| *Bedridden 7 days or more* | 6 (5.5%) | 12 (4.2%) | 17 (3.4%) |
| Feeling your heart pound or race - Bothered a lot |  |  |  |
| non-COVID group (%) | 1077 (5.0%) | 646 (2.6%) | 191 (1.5%) |
| COVID-19 group (%) | 164 (10.5%) | 183 (3.9%) | 357 (2.2%) |
| 0-2 months |  |  |  |
| *Not bedridden* | 2 (2.9%) | 14 (3.3%) | 7 (1.2%) |
| *Bedridden 1-6 days* | 11 (15.9%) | 24 (4.9%) | <5 |
| *Bedridden 7 days or more* | 9 (18.4%) | 5 (6.9%) | 7 (12.5%) |
| 3-5 months |  |  |  |
| *Not bedridden* | 17 (6.6%) | 4 (3.5%) | 137 (1.8%) |
| *Bedridden 1-6 days* | 36 (13.1%) | 3 (2.9%) | 31 (3.1%) |
| *Bedridden 7 days or more* | 30 (21.7%) | 3 (5.2%) | 30 (5.7%) |
| 6-9 months |  |  |  |
| *Not bedridden* | 10 (7.5%) | 5 (2.5%) | 49 (1.5%) |
| *Bedridden 1-6 days* | 13 (8.4%) | 3 (1.5%) | 14 (3.1%) |
| *Bedridden 7 days or more* | 2 (3.8%) | 10 (8.1%) | 12 (3.8%) |
| 10-27 months |  |  |  |
| *Not bedridden* | 3 (2.2%) | 12 (3.3%) | 29 (1.8%) |
| *Bedridden 1-6 days* | 16 (14.3%) | 7 (2.1%) | <5 |
| *Bedridden 7 days or more* | 14 (12.7%) | 26 (9.1%) | 34 (6.8%) |
| Shortness of breath - Bothered a lot |  |  |  |
| non-COVID group (%) | 518 (2.4%) | 414 (1.7%) | 173 (1.4%) |
| COVID-19 group (%) | 89 (5.7%) | 155 (3.3%) | 206 (1.3%) |
| 0-2 months |  |  |  |
| *Not bedridden* | 3 (4.3%) | 10 (2.4%) | 8 (1.3%) |
| *Bedridden 1-6 days* | 6 (8.7%) | 16 (3.3%) | <5 |
| *Bedridden 7 days or more* | 6 (12.2%) | 8 (11.1%) | <5 |
| 3-5 months |  |  |  |
| *Not bedridden* | 8 (3.1%) | 1 (0.9%) | 69 (0.9%) |
| *Bedridden 1-6 days* | 16 (5.8%) | 3 (2.9%) | 14 (1.4%) |
| *Bedridden 7 days or more* | 14 (10.1%) | 6 (10.3%) | 17 (3.2%) |
| 6-9 months |  |  |  |
| *Not bedridden* | 3 (2.2%) | 7 (3.6%) | 28 (0.9%) |
| *Bedridden 1-6 days* | 9 (5.8%) | 4 (2.0%) | <5 |
| *Bedridden 7 days or more* | 3 (5.8%) | 7 (5.7%) | 13 (4.1%) |
| 10-27 months |  |  |  |
| *Not bedridden* | 5 (3.6%) | 5 (1.4%) | 18 (1.1%) |
| *Bedridden 1-6 days* | 4 (3.6%) | 5 (1.5%) | <5 |
| *Bedridden 7 days or more* | 11 (10.0%) | 21 (7.4%) | 27 (5.4%) |

^a^ Prevalence for number less than 5 was not reported in DBDS due to the data regulation in Denmark.

**Supplementary figure**

**Figure S1: Flowchart for study with all cohorts and overall participation**

64 880 participants included in analysis from all cohorts

14 358 participants

18 190 participants

3 310 participants

C-19 Resilience (IS)

N=14 822

Omtanke2020 (SE)

N= 18 759

MAP-19 (NO)

N=3 310

65 913 participants in all cohorts and received questionnaires on PHQ-15

Excluded due to missing values on:

1.COVID-19 diagnosis: 237

2.physical symptom: 227^*^

Excluded due to missing values on:

1.COVID-19 diagnosis: 0

2.physical symptom: 569^*^

Excluded due to missing values on:

1.COVID-19 diagnosis: 0

2.physical symptom: 0

DBDS (DK)

N=29 022

29 022 participants

Excluded due to missing values on:

1.COVID-19 diagnosis: 0

2.physical symptom: 0

*Participants were excluded due to missingness on more than 4 out of 15 items of PHQ-15 questionnaire. Multiple imputation was used to fulfill the missing options on PHQ-15 score for remaining participants.

### **Figure S2. Prevalence ratio (95% confidence interval) of severe physical symptoms (PHQ-15≥15) among persons with COVID-19 compared with individuals *NOT* diagnosed with COVID-19, by current mental health indicators (current depressive, anxiety, and COVID-19 related distress symtoms) and pre-existing comorbidity.^a^**

A. **A meta analyses on all cohorts**

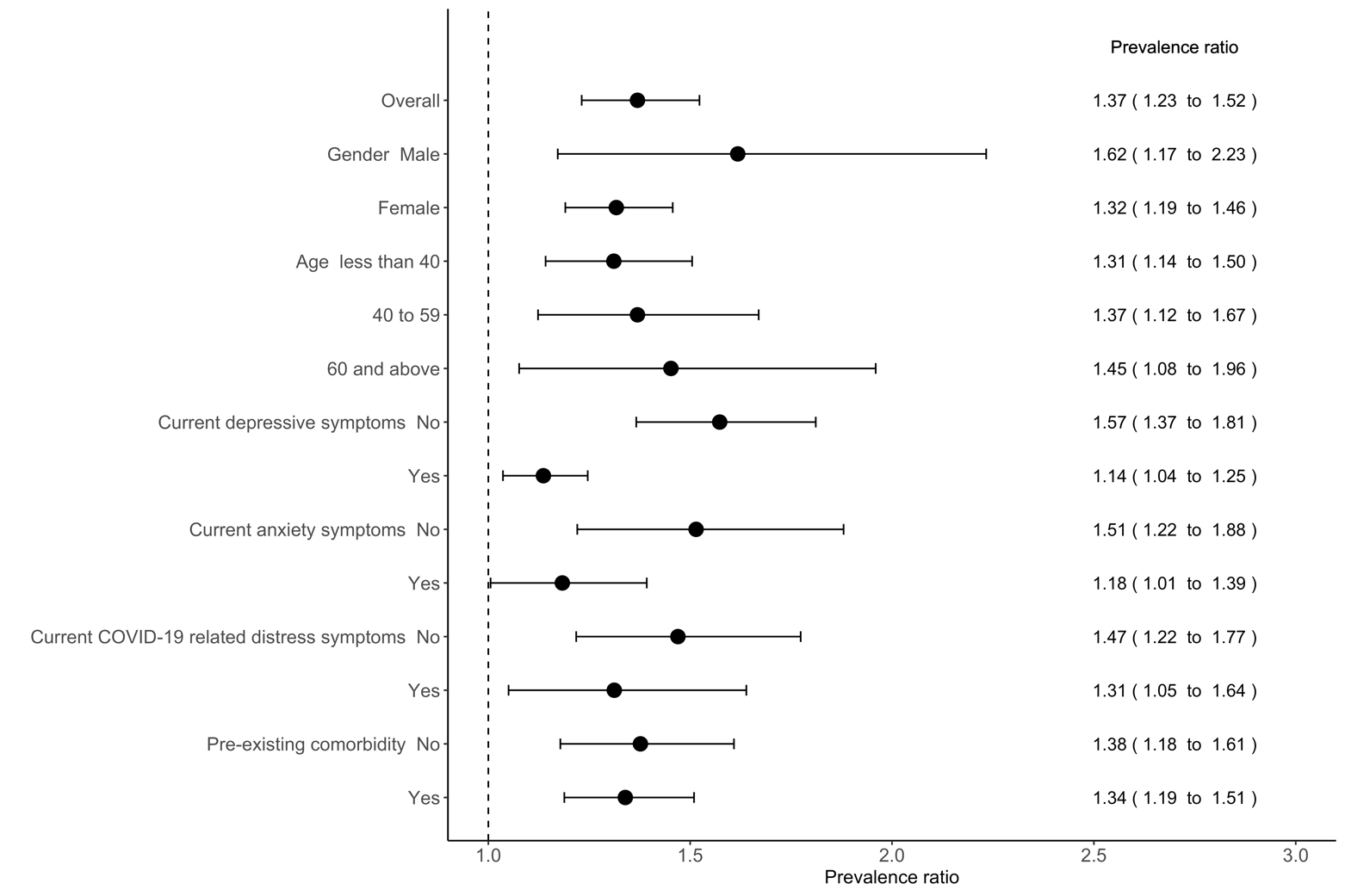

B. **By each cohort**

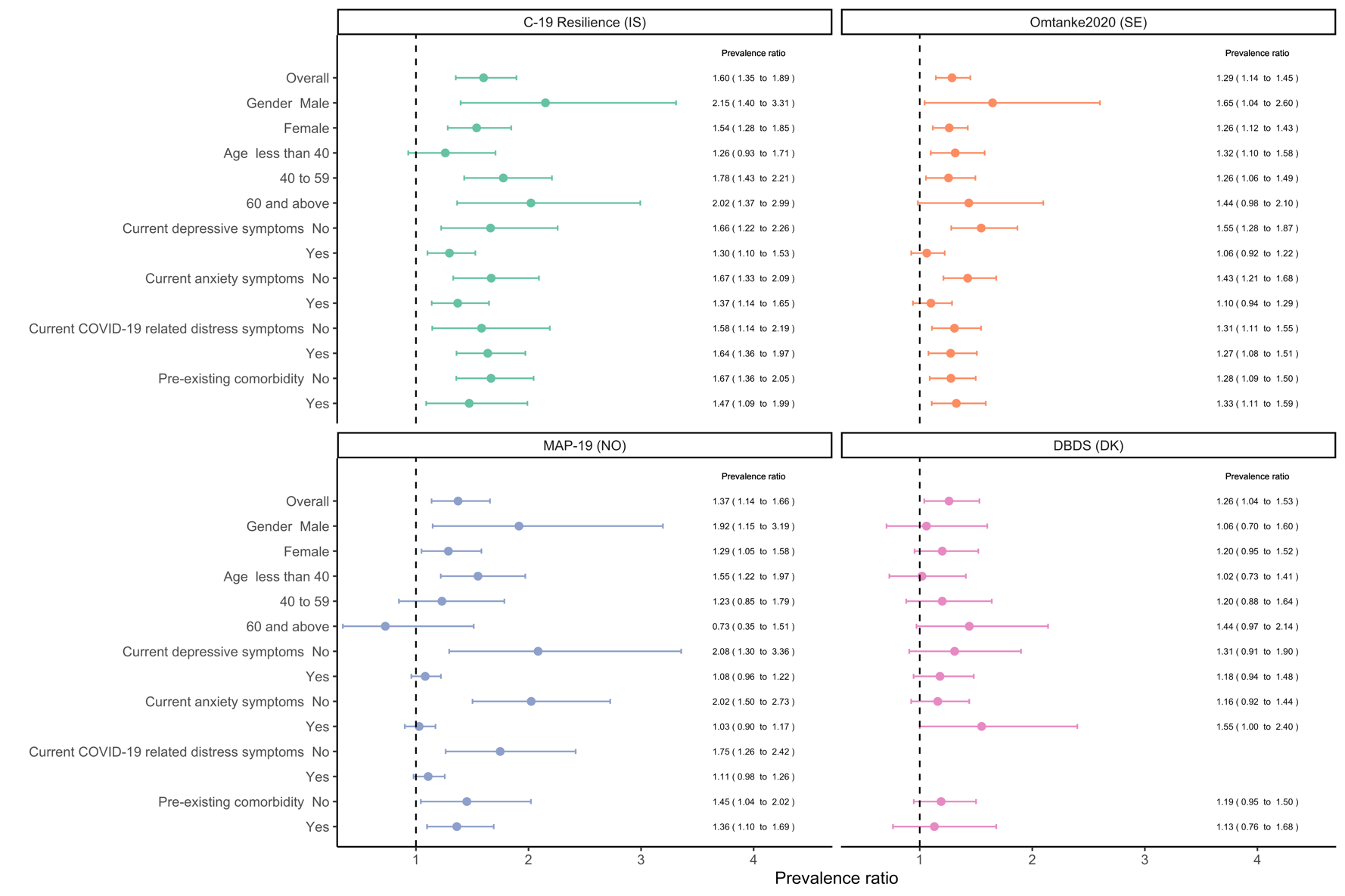

**^a^** Prevalence ratios were adjusted for age, gender, residency, average monthly income, current smoking, BMI, pre-existing comorbidity, relationship status, habitual drinking, previous diagnosis of psychiatric disorder, and response period in overall association. Income was not available in Omtanke2020 (SE), and relationship status and current COVID-19 realted distress symptoms were not available in DBDS (DK).

### **Figure S3. Prevalence ratio (95% confidence interval) of severe physical symptoms (PHQ-15≥15) among individuals with COVID-19 compared with individuals *NOT* diagnosed with COVID-19 in each cohort, by illness severity according to time since diagnosis^a^.**

A. By illness severity – bedridden

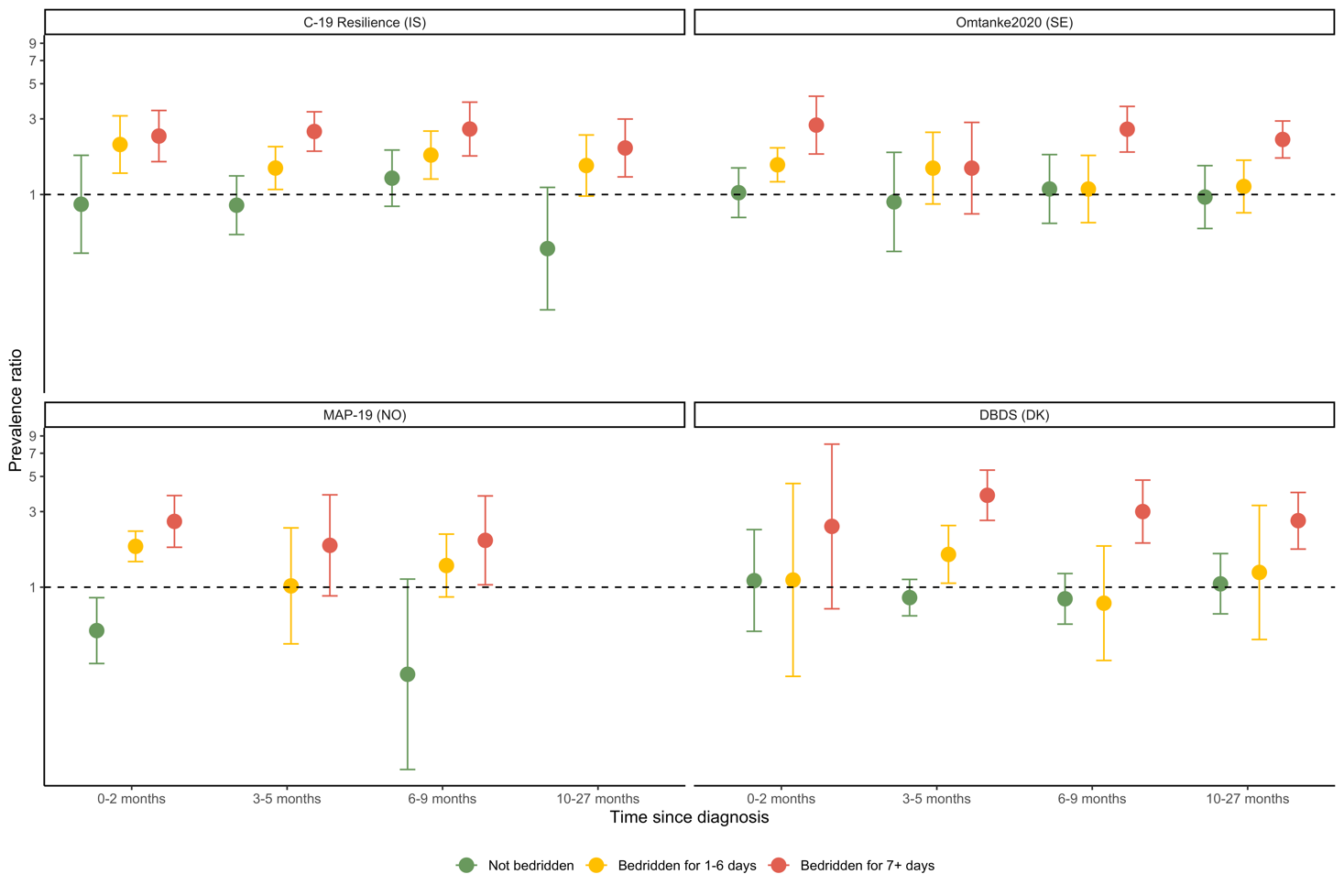

B. By illness severity – hospitalization (C-19 Resilience and Omtanke2020)

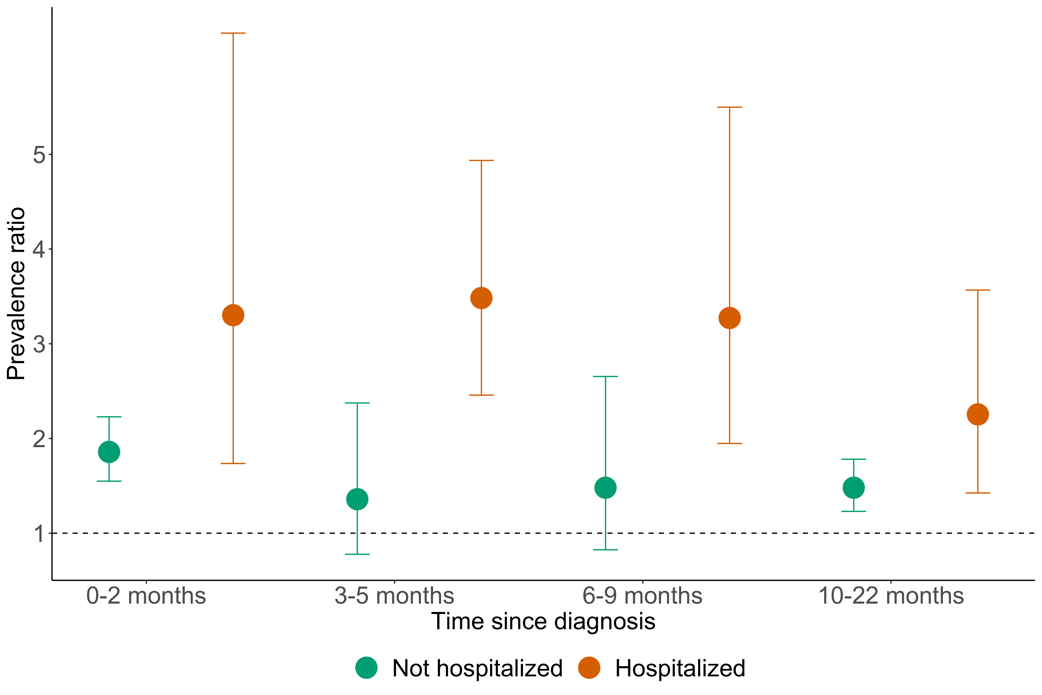

**^a^** Individuals with missing information on time since diagnosis or illness severity were excluded from this analysis. Prevalence ratios were adjusted for age, gender, residency, average monthly income, current smoking, BMI, pre-existing comorbidity, relationship status, habitual drinking, previous diagnosis of psychiatric disorder, and response period. Income was not available in Omtanke2020 (SE), and relationship status was not available in DBDS (DK).

### **Figure S4. Prevalence ratios (95% confidence interval) of physical symptom severity among individuals with COVID-19 compared with individuals *NOT* diagnosed with COVID-19 in each cohort, by individual physical symtom^a^.**

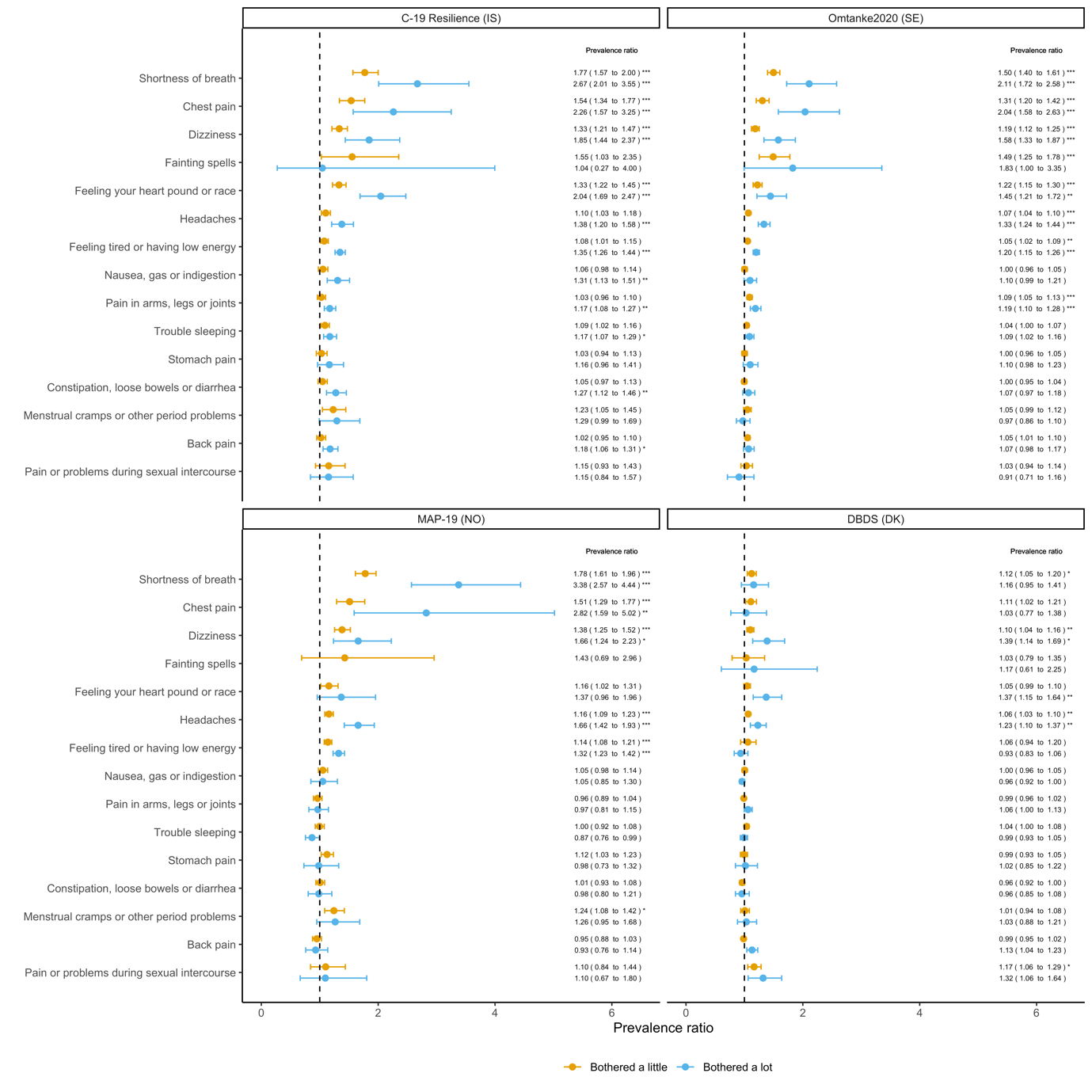

**^a^** Prevalence ratios were adjusted for age, gender, residency, average monthly income, current smoking, BMI, pre-existing comorbidity, relationship status, habitual drinking, previous diagnosis of psychiatric disorder, and response period. Income was not available in Omtanke2020 (SE), and relationship status was not available in DBDS (DK). Menstrual cramps were only applied to women aged <60 years. P-values were corrected for multiple testing using Bonferroni correction method. * indicates corrected P-value<0.05; **<0.01; ***<0.001.

**Figure S5. Prevalence ratios (95% confidence interval) of reporting *bothered a lot* to each symptom among individuals with COVID-19 compared with individuals *NOT* diagnosed with COVID-19 in C-19 Resilience and Omtanke2020, by illness severity (hospitalization) according to time since diagnosis^a^**

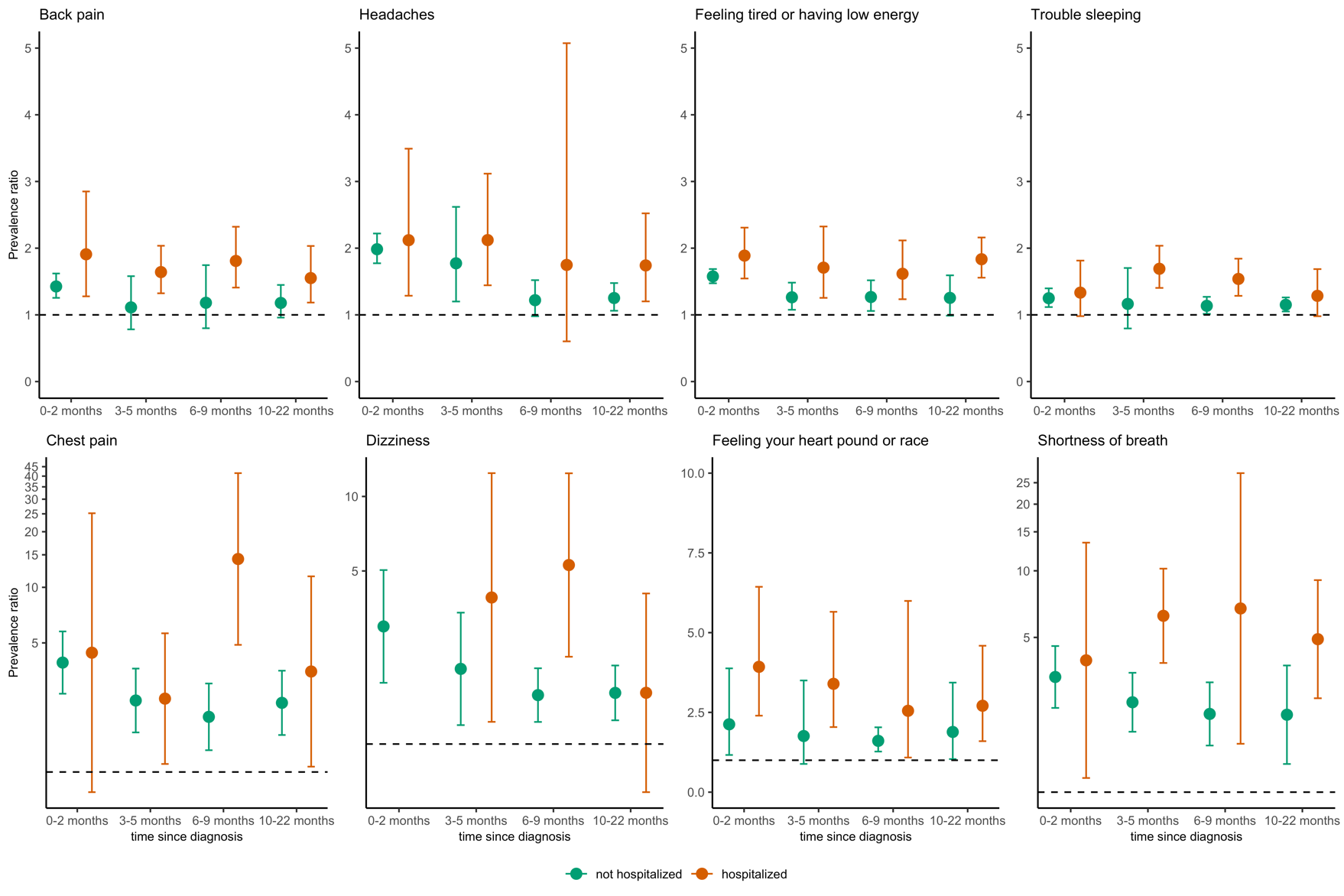

**^a^** Prevalence ratios were adjusted for age,gender, residency, average monthly income, current smoking, BMI, pre-existing comorbidity, relationship status, habitual drinking, previous diagnosis of psychiatric disorder, and response period. Income was not available in Omtanke2020 (SE).
